# COVID-HEART: Development and Validation of a Multi-Variable Model for Real-Time Prediction of Cardiovascular Complications in Hospitalized Patients with COVID-19

**DOI:** 10.1101/2021.01.03.21249182

**Authors:** Julie K. Shade, Ashish N. Doshi, Eric Sung, Dan M. Popescu, Anum S. Minhas, Nisha A. Gilotra, Konstantinos N. Aronis, Allison G. Hays, Natalia A. Trayanova

## Abstract

Cardiovascular (CV) manifestations of COVID-19 infection carry significant morbidity and mortality. Current risk prediction for CV complications in COVID-19 is limited and existing approaches fail to account for the dynamic course of the disease. Here, we develop and validate the COVID-HEART predictor, a novel continuously-updating risk prediction technology to forecast CV complications in hospitalized patients with COVID-19. The risk predictor is trained and tested with retrospective registry data from 2178 patients to predict two outcomes: cardiac arrest and imaging-confirmed thromboembolic events. In repeating model validation many times, we show that it predicts cardiac arrest with an average median early warning time of 18 hours (IQR: 13-20 hours) and an AUROC of 0.92 (95% CI: 0.91-0.92), and thromboembolic events with a median early warning time of 72 hours (IQR: 12-204 hours) and an AUROC of 0.70 (95% CI: 0.67-0.73). The COVID-HEART predictor is anticipated to provide tangible clinical decision support in triaging patients and optimizing resource utilization, with its clinical utility potentially extending well beyond COVID-19.

## Main

Patients with COVID-19, the disease caused by the novel severe acute respiratory syndrome coronavirus 2 (SARS-CoV-2), often present with cardiovascular (CV) manifestations such as myocardial infarction, thromboembolism, and heart failure.^1^ Clinically overt cardiac injury or cardiomyopathy is reported in 8 to 33% of hospitalized patients^2,3^ and is associated with up to 50% mortality,^4^ but imaging studies suggest the true incidence of cardiac involvement in all persons infected with SARS-CoV-2 could be as high as 60%.^5^ Thromboembolic events are also frequently reported in severe COVID-19 and are associated with mortality; one study found that 70.1% of non-survivors and 0.6% of survivors met criteria for disseminated intravenous coagulation.^6^ Furthermore, thromboembolic complications are more pronounced in acute COVID-19 infection than in other viral illnesses, and include pulmonary embolus and ischemic stroke, which can be fatal and are a significant cause of morbidity even as the infection resolves.^7^ Despite the prevalence of thromboembolism and cardiac injury and their associations with poor outcomes,^6,8,9^ no approach currently exists to forecast adverse CV events in COVID-19 patients in real time.

Machine learning (ML) techniques are ideal for discovering patterns in high-dimensional biomedical data, especially when little is known about the underlying biophysical processes. ML is thus well-positioned for applications in COVID-19 and indeed has been employed in screening, contract tracing, drug development, and outbreak forecasting.^10,11^ ML approaches have been developed for prognostic assessment of hospitalized patients with COVID-19, including models which predict in-hospital mortality,^12–17^ progression to severe disease,^14,18–21^ and outcomes related to respiratory function.^10,15,22^ A model was also proposed for prediction of thromboembolic events but it required that all variables be present for all patients, did not provide dynamic risk updates, and was trained with data from only 76 patients.^23^ Thus far, prognostic models have relied on clinical data available at a single time-point,^10,22^ and have not accounted for the dynamic and difficult-to-predict course of this novel disease.

Here, we develop and validate a prognostic ML model to forecast the real-time risk of CV complications in hospitalized patients with COVID-19. We term the model the COVID-HEART predictor. We focus on predicting two clinically important CV outcomes in COVID-19, in-hospital cardiac arrest and thromboembolic events. In-hospital cardiac arrest is a clearly identifiable outcome and is often CV-related, thus it was selected to demonstrate the potential utility of the COVID-HEART predictor. Thromboembolic events are more difficult to identify and require imaging confirmation, thus, this outcome was selected to demonstrate the versatility of the COVID-HEART predictor in analyzing real-world clinical data and handling CV-specific outcomes. The model is trained and tested with data from over 2000 patients with SARS-CoV-2 infection, confirmed by positive polymerase chain reaction or nucleic acid testing, admitted to multiple hospitals within a single health system. The resulting risk predictor is robust to missing data and can be updated each time new data becomes available, representing a continuously evolving warning system for an impending event. It can also predict the likelihood of an adverse event within multiple timeframes (e.g. 2 hours, 8 hours, 24 hours). The COVID-HEART predictor is anticipated to be of great clinical use in triaging patients and optimizing resource utilization by identifying at-risk patients in real time.

Fig.1 presents a schematic of the COVID-HEART continuously-updating risk prediction technology. The TRIPOD guidelines for development, validation, and presentation of a multivariable prediction model,^24^ as recommended by Wyants et al,^22^ were followed here (Supplementary Table S1). The model uses features extracted from 106 different clinical data inputs, some of which are associated with CV complications in COVID-19 and in other severe respiratory illnesses (Supplementary Table S2). To avoid bias, variables that were directly impacted by a physician’s assessment of the patient’s condition, such as the fraction of inspired oxygen set on a mechanical ventilator, are excluded. The COVID-HEART predictor is trained to estimate the probability that a patient will experience a particular CV event within a set number of hours (outcome window) after any point during the patient’s hospitalization. It uses static variables (demographics and comorbidities) and dynamic clinical data collected during time periods of markedly different duration prior to the time point of prediction: data collected just prior (short features), data collected over the entire hospitalization (long features), and data collected over the entire hospitalization weighted such that recent data is assigned higher importance (exponentially weighted decaying features).

**Figure 1:**
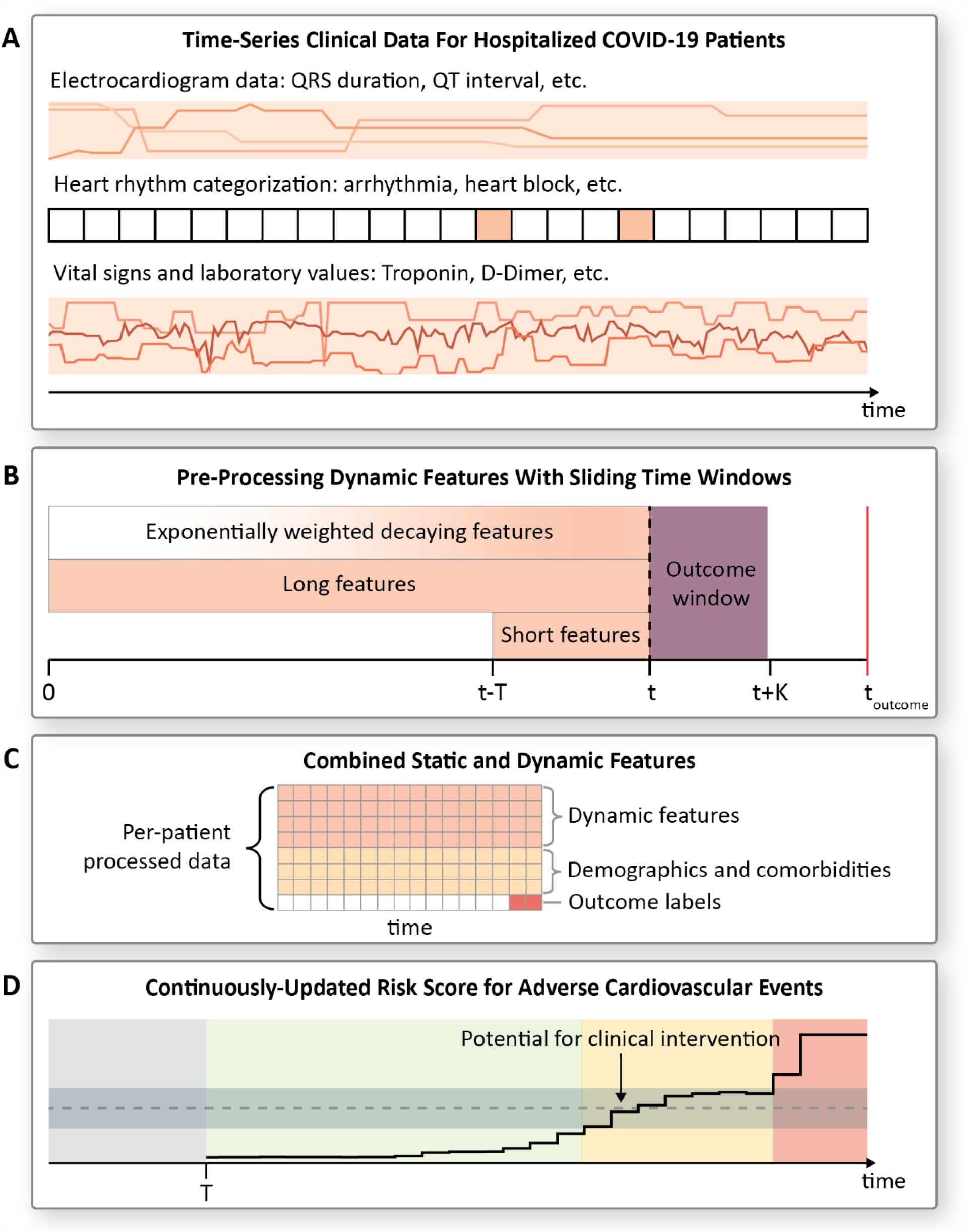
Schematic Overview of COVID-HEART Study. **(A)** Time-series clinical data used as input. The continuously-updating COVID-HEART predictor uses features extracted from 106 clinical data inputs including electrocardiogram data, heart rhythm categorizations, vital signs, laboratory values, demographics, and comorbidities. Supplementary Table S3 provides a complete list of inputs. Data shown here are representative and do not correspond with the risk score shown in (D); Fig.3 provides examples of real data and resulting risk scores over time for patients in the test set for the two cardiovascular (CV) outcomes: cardiac arrest and thromboembolic events. Electrocardiogram (ECG) data consists of measured parameters from a 12-lead ECG (QRS duration, QT interval, etc.), these are recorded about once per day per patient. Heart rhythm categorizations and vital signs are extracted from flowsheet data. Heart rhythm annotations (e.g. atrial fibrillation, ventricular tachycardia, heart block) are processed with a binary label indicating whether a patient experienced each heart rhythm during a given time window, and an integer reflecting how many times it was recorded. Colored blocks represent time-windows in which a given heart rhythm was recorded. Laboratory values include variables which have been shown to be associated with adverse cardiovascular outcomes in COVID-19, such as D-Dimer and lymphocytes, and standard lab tests such as sodium and potassium levels. Each clinical measurement is assumed constant until a new value is recorded. Patients with missing data were not excluded to ensure the model was robust to normal variations in the quantity and quality of clinical data collected in a real-world setting. **(B)** Dynamics features pre-processing with sliding time windows. The dynamic (time-series) features are pre-processed for each patient with sliding time windows; at each time point, features are recorded for the window prior (“feature window”) and associated with a binary label (0,1) indicating whether the patient experienced a given outcome in the following K hours (“outcome window”). The classifier uses features calculated from clinical data at multiple time durations: “short features”, which encompass a short period prior to the time point in question, and “long features” and “exponentially weighted decaying features”, which encompass the patient’s entire hospitalization. The relative intensity levels within the three feature windows represent the weighting of values at each time; darker colors indicate higher weight. The features derived from the dynamic clinical inputs include the mean, minimum, maximum, standard deviation, and amplitude of first frequency in Fourier space. Data is censored at the time of discharge or at the time of outcome, whichever comes first. The COVID-HEART predictor is trained to predict in-hospital events only. **(C)** Combined features. For each time window, the processed dynamic features are combined with static features including demographics and comorbidities. Table 1 lists the most important features for prediction of each outcome. The model is a linear classifier trained with stochastic gradient descent; this training approach allows the model to be updated with new data without needing full re-training. Pre-processing steps include dropping features that are missing for more than 60% of time windows in the training set, mean-value imputation of remaining missing values, standardization, and feature selection using a random-forest to minimize multi-collinearity in the selected features. Each time window is treated as a separate data point during training, but all the time windows from each patient are assigned to the same fold of stratified 5-fold cross-validation. Outcome labels are per-window, so for a patient that experienced an event all time windows would be labeled as “no outcome” except for the windows immediately before the event. In a secondary analysis, we investigated whether a larger proportion of time windows were predicted positive for patients who eventually experienced the outcome than for patients who did not. Detail is provided in Methods. **(D)** Continuously-updating risk score. The COVID-heart predictor is trained to provide a binary output indicating whether the patient is at-risk for an adverse CV event (e.g. cardiac arrest, thromboembolic event) in the K hours following a given time point, then is calibrated to provide a risk score (probability) for the outcome. Shown is a sample risk score for a patient that experienced an event: green color indicates low risk score; yellow indicates a risk score within a pre-determined range of a threshold value, and the red indicates that the patient is at high risk for an event in the following K hours. The gray color represents the first T hours of the patient’s hospitalization, during which data is being collected to inform the initial risk score for the patient. T is the duration of the short feature window. We envision that clinical intervention could be made in the “yellow zone” to avert the impending event.

The COVID-HEART predictor was trained with stochastic gradient descent; the training approach allows, in practice, for continuous model update with new data without full re-training, thus ensuring accurate risk predictions as COVID-19 treatment paradigms evolve. Three configurations of a linear classifier were investigated: one with “short features” only, one with features from multiple time durations, and one with features from multiple time durations that used, during training, the Synthetic Minority Oversampling TEchnique (SMOTE) for nominal and continuous variables^25^ and random undersampling of the majority class to account for severe class imbalance. During training, losses were weighted to strongly penalize prediction errors for “positive” time windows (time windows for which the patient experienced the CV outcome in the following outcome window). to further account for class imbalance, and the optimal classifier configuration for prediction of each outcome was selected based on the cross-validation area under the receiver operating characteristic curve (AUROC) following probability calibration. The training process and usage of the predictor are described in detail in Methods.

**Table 1:** Up to 20 features with largest coefficients for prediction of cardiac arrest and thromboembolic events. In the table, “Feature” refers to the processed input to the ML algorithm based on the values of each clinical data input during each time window, and “Time Duration” refers to the length of time over which clinical data values were considered to calculate each feature. Note that all features were normalized with a mean of 0 and a standard deviation of 1 during pre-processing, although raw values are shown here, and that values are listed per time-window, not per-patient. The coefficients are applied after normalization. The optimal loss functions were log and modified Huber for prediction of cardiac arrest and thromboembolic events, respectively, and the classifiers were calibrated, so coefficient interpretation is non-trivial especially for prediction of thromboembolic events. These are not the only features included in the classifier for prediction of cardiac arrest. There were only 3 features included in the classifier for prediction of thromboembolic events due to the low number of events in the development set. P-values calculated using two-sample two-sided t-test or chi-squared test as appropriate. This table was generated using the python package *tableone*.^30^ Comorbidities, including chronic lung disease and pulmonary circulation disorders, are defined using ICD-10 codes according to the Elixhauser comorbidity definitions.^31^ *Abbreviations: white blood cell count (WBC), diastolic blood pressure (DBP), systolic blood pressure (SBP), glomerular filtration rate (GFR), blood urea nitrogen (BUN), c-reactive protein (CRP), activated partial thromboplastin time (aPTT), exponentially weighted decaying (Exp. Decay), coefficient (coef)*.

|  |  |  |  | Train - Cardiac Arrest |  |  |  | Test - Cardiac Arrest |  |  |  |
| --- | --- | --- | --- | --- | --- | --- | --- | --- | --- | --- | --- |
| Input | Feature | Duration | Coef | Missing (%) | No Outcome | Outcome | P | Missing (%) | No Outcome | Outcome | P |
|  |  |  |  |  | n = 412198 | n = 440 |  |  | n = 101510 | n = 109 |  |
| Chronic Lung Disease |  | Static | 0.318 | 0.001 | 85419 (20.8) | 98 (22.3) | 0.475 | 0.000 | 21449 (21.2) | 34 (31.2) | 0.014 |
| Pulse | Maximum | 2 h | 0.308 | 0.098 | 87.7 (17.1) | 104.5 (23.4) | <0.001 | 0.072 | 87.1 (17.0) | 106.1 (22.5) | <0.001 |
| White Blood Cell Count | Minimum | 2 h | 0.268 | 0.074 | 9.4 (5.0) | 14.7 (7.8) | <0.001 | 0.081 | 9.0 (4.6) | 14.3 (8.0) | <0.001 |
| Fibrinogen | Maximum | 2 h | 0.242 | 0.099 | 21.9 (6.1) | 27.1 (8.3) | <0.001 | 0.074 | 22.0 (6.2) | 26.8 (9.2) | <0.001 |
| Pulse O2 Sat | Mean | 2 h | -0.215 | 0.099 | 95.8 (2.8) | 90.4 (7.1) | <0.001 | 0.073 | 95.8 (2.7) | 91.8 (6.8) | <0.001 |
| DBP | Maximum | 2 h | -0.204 | 0.098 | 70.1 (12.5) | 57.7 (14.3) | <0.001 | 0.072 | 69.0 (12.0) | 57.1 (16.1) | <0.001 |
| Pulse O2 Sat | Maximum | 2 h | -0.199 | 0.099 | 96.4 (2.7) | 91.2 (7.2) | <0.001 | 0.073 | 96.4 (2.6) | 92.6 (7.0) | <0.001 |
| SBP | Maximum | 2 h | -0.193 | 0.098 | 129.6 (21.3) | 108.6 (27.8) | <0.001 | 0.072 | 128.9 (21.4) | 112.6 (30.6) | <0.001 |
| SBP | Minimum | 2 h | -0.175 | 0.098 | 121.3 (20.3) | 99.7 (26.6) | <0.001 | 0.072 | 120.4 (19.8) | 100.6 (30.2) | <0.001 |
| GFR | Maximum | 2 h | -0.168 | 0.066 | 79.8 (38.5) | 52.4 (36.2) | <0.001 | 0.071 | 83.6 (39.4) | 60.1 (39.3) | <0.001 |
| Sodium | Minimum | 2 h | 0.159 | 0.067 | 139.5 (5.2) | 142.6 (6.9) | <0.001 | 0.072 | 139.3 (5.3) | 143.4 (8.5) | <0.001 |
| Platelet Count | Maximum | 2 h | -0.155 | 0.073 | 275.8 (136.0) | 215.3 (125.1) | <0.001 | 0.082 | 294.5 (153.3) | 231.9 (133.7) | <0.001 |
| BUN | Minimum | 2 h | 0.153 | 0.068 | 25.7 (19.3) | 44.1 (30.3) | <0.001 | 0.072 | 24.1 (18.2) | 38.3 (25.4) | <0.001 |
| Temperature | St Dev | 2 h | -0.153 | 0.102 | 0.1 (0.2) | 0.1 (0.1) | <0.001 | 0.077 | 0.1 (0.2) | 0.1 (0.2) | 0.771 |
| Anion Gap | Minimum | 2 h | 0.143 | 0.067 | 12.0 (4.1) | 15.4 (5.8) | <0.001 | 0.072 | 11.9 (4.0) | 14.7 (5.6) | <0.001 |
| Lactate | Minimum | 2 h | 0.137 | 0.595 | 1.4 (0.7) | 4.1 (4.2) | <0.001 | 0.612 | 1.5 (0.7) | 4.5 (3.9) | <0.001 |
| Pulse | St Dev | 2 h | 0.112 | 0.098 | 2.1 (3.5) | 3.5 (6.4) | <0.001 | 0.072 | 2.1 (3.4) | 3.3 (5.4) | 0.025 |
| CRP | Maximum | 2 h | 0.106 | 0.202 | 26.7 (49.0) | 63.7 (80.6) | <0.001 | 0.198 | 24.9 (45.4) | 68.9 (84.6) | <0.001 |
| Chloride | Minimum | 2 h | 0.105 | 0.067 | 102.6 (5.9) | 104.9 (7.5) | <0.001 | 0.072 | 102.0 (5.8) | 105.2 (9.1) | 0.001 |
| Normal Sinus Rhythm | Maximum | 2 h | -0.104 | 0.098 | 0.1 (0.3) | 0.1 (0.2) | <0.001 | 0.072 | 0.1 (0.3) | 0.0 (0.2) | <0.001 |
|  |  |  |  | Train – Thromboembolic Event |  |  |  | Test – Thromboembolic Event |  |  |  |
| Input | Feature | Duration | Coef | Missing (%) | No Outcome | Outcome | P | Missing (%) | No Outcome | Outcome | P |
|  |  |  |  |  | n = 14854 | n = 26 |  |  | n = 3514 | n = 6 |  |
| QRS Duration | St Dev | Long | 0.291 | 0.207 | 3.6 (0.5) | 3.4 (0.4) | 0.011 | 0.207 | 3.7 (0.5) | 3.8 (0.3) | 0.570 |
| Platelet Count | Maximum | Long | 0.059 | 0.086 | 8.4 (2.6) | 9.0 (2.4) | 0.277 | 0.065 | 8.5 (2.7) | 9.0 (2.8) | 0.679 |
| aPTT | St Dev | 24 h | 0.012 | 0.109 | 75.5 (14.2) | 78.6 (16.0) | 0.362 | 0.086 | 76.1 (15.1) | 85.3 (12.1) | 0.121 |

The COVID-HEART predictor was developed and validated in a retrospective study of patients with confirmed SARS-CoV-2 infection admitted to any one of the 5 Johns Hopkins Health system hospitals between March 1, 2020 and September 27, 2020. Supplementary Fig.S1 shows the flow of patients through the study. In-hospital cardiac arrest and in-hospital thromboembolic events were predicted on a continuous basis; outcome definition is detailed in Methods. 2178 patients met eligibility criteria for cardiac arrest prediction, of whom 277 (12.7%) experienced in-hospital cardiac arrest. 1601 patients met eligibility criteria for thromboembolic event prediction, of whom 32 (2.0%) experienced imaging-confirmed in-hospital thromboembolic events. Patients were divided into development (80%) and test (20%) sets with stratified random selection. Supplementary Tables S3 and S4 provide demographic and clinical comparisons between patients who did and did not experience each outcome, and between the training and test sets.

COVID-HEART performance for the two CV outcomes, in-hospital cardiac arrest and thromboembolic events, is summarized in Fig.2. The figure presents five-fold stratified patient-based cross-validation and test performance results for each of the three different classifier configurations. The optimal classifier configurations were selected based on the cross-validation area under the receiver operating characteristic curve (AUROC). Following the initial train-test split, the results of which were presented in Fig.2 and investigated in detail in Fig.3 and Table 1, an “outer loop” of cross-validation was added and results over multiple iterations were aggregated to obtain 95% confidence intervals for the cross-validation and testing AUROCs (Fig.2B-C). The mean cross-validation and test AUROCs were 0.91 (95% CI: 0.91-0.92) and 0.92 (95% CI: 0.91-0.92) for prediction of cardiac arrest and 0.77 (95% CI: 0.75-0.79) and 0.70 (95% CI: 0.67-0.73) for prediction of thromboembolic events, respectively.

**Figure 2:**
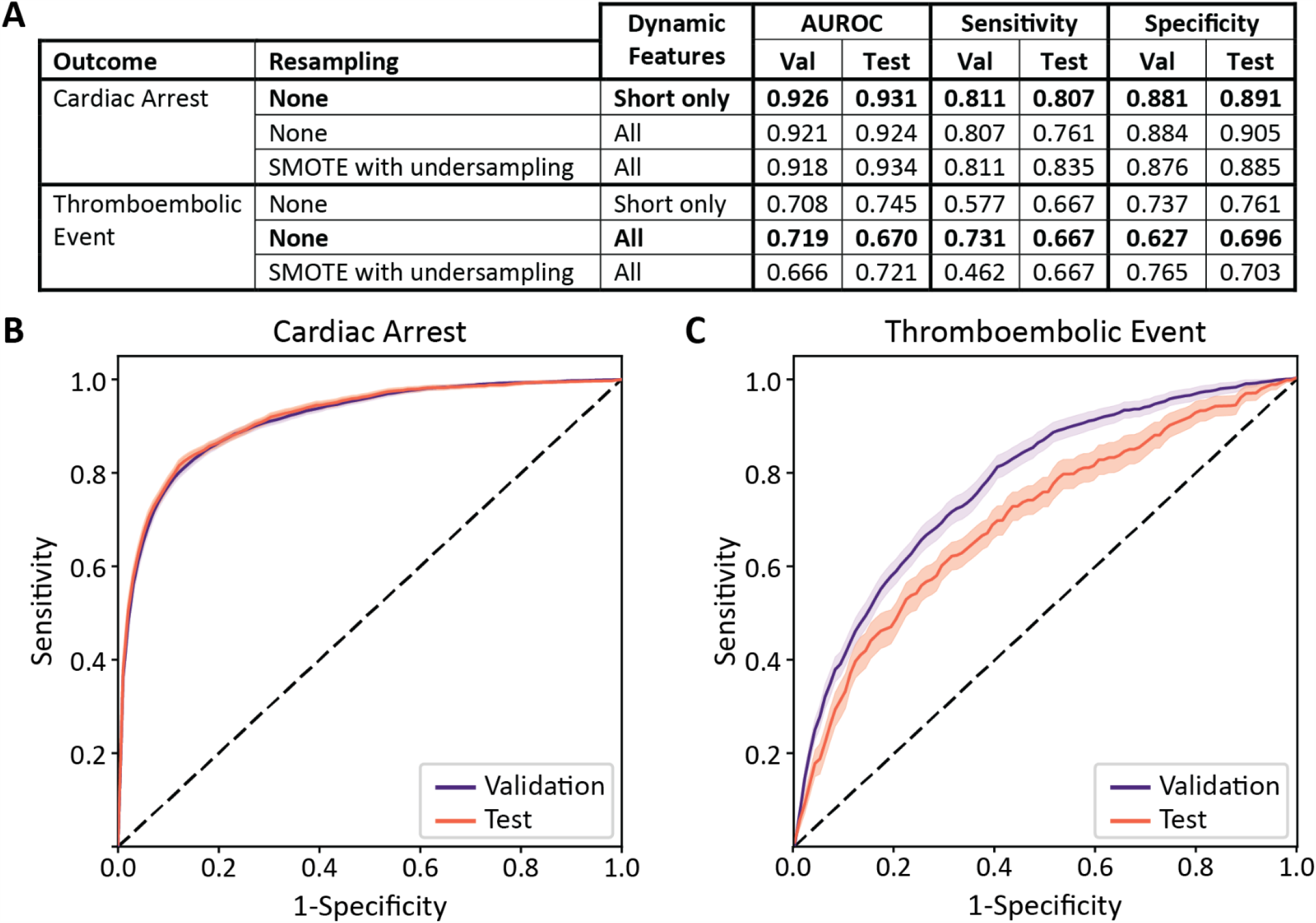
The COVID-HEART predictor can accurately predict the risk of cardiac arrest and thromboembolic events in real time. **(A) COVID-HEART** performance metrics for the two CV outcomes: cardiac arrest and thromboembolic events. Shown are 5-fold stratified patient-based cross-validation and test performance metrics for three linear classifier configurations. Cardiac arrest predictions presented here are for an outcome window of 2 hours, short-time feature window of 2 hours, and time-step of 1 hour. Thromboembolic event predictions shown here are for an outcome window of 24 hours, short-time feature window of 24 hours, and time-step of 24 hours. Using different time-steps and feature and outcome windows for predicting the different outcomes is necessitated by the granularity of the outcomes within the electronic health record; thromboembolic events require imaging confirmation and are thus are only recorded on the date they occurred, while cardiac arrest is recorded with the exact time. The best-performing of the three linear classifier models for prediction of each CV outcome are bolded. These were selected based on the area under the receiver operating characteristic curve (AUROC). **(B)** Risk of cardiac arrest prediction. Cross-validation (purple) and testing (orange) receiver operating characteristic (ROC) curves for prediction of cardiac arrest using the optimal classifier configuration: a linear classifier with no resampling and short-term features only. To generate the ROC curves, 6 full iterations of 5-fold nested patient-based cross validation were run resulting in a total of 30 test sets and 150 inner loops of cross-validation. Shaded regions represent the 95% confidence interval of each ROC curve. **(C)** Risk of thromboembolic event prediction. Cross-validation (purple) and testing (orange) receiver operating characteristic (ROC) curves for prediction of thromboembolic events using optimal classifier configuration: a linear classifier with no resampling and all feature types. To generate the ROC curves, 8 full iterations of 5-fold nested patient-based cross validation were run resulting in a total of 40 test sets and 200 inner loops of cross-validation. Shaded regions represent the 95% confidence interval of each ROC curve.

**Figure 3:**
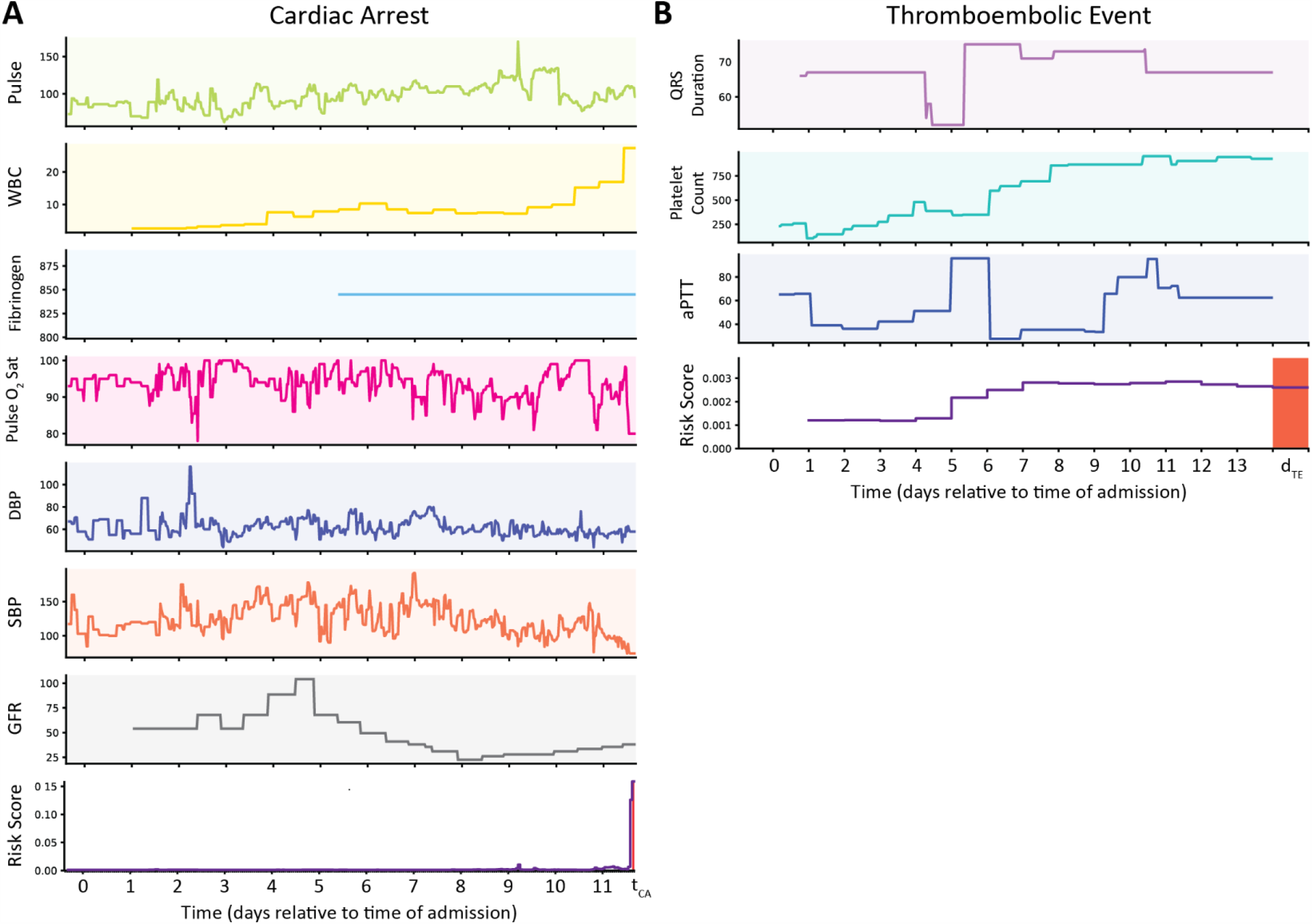
Two examples of “true positive” predictions for two different patients, one from the cardiac arrest test set and one from the thromboembolic event test set, using the COVID-HEART predictor. **(A)** Clinical time-series inputs (top 7 rows) from which the features with the largest coefficients were derived for prediction of cardiac arrest, and time-series risk score (bottom row) for a patient who experienced cardiac arrest during their hospitalization, and for whom the classifier’s prediction was correct prior to the cardiac arrest. The most important features derived from these inputs are listed in Table 1. A new prediction is generated every hour. The risk score is low for the first 11 days of the patient’s admission, then rapidly increases in the 2 hours preceding the cardiac arrest. The date refers to the days of admission relative to midnight on the first full day of admission. Units for each predictor are as follows: Pulse (beats/minutes), WBC (cells/ mm^3^), Fibrinogen (mg/dL), Pulse O_2_ saturation (%), DBP (mmHg), SBP (mmHg), GFR (mL/min) **(B)** Clinical time-series inputs (top 3 rows) from which the 3 selected features were derived for prediction of thromboembolic events, and time-series risk score (bottom row) for a patient who experienced a thromboembolic event during their hospitalization. The most important features derived from these inputs are listed in Table 1. The patient’s risk score increases from days 5-7 and then remains elevated leading up to the thromboembolic event, which occurs on day 14 of the admission. The binary risk threshold is 0.002, selected automatically to balance sensitivity and specificity for the development data. The x-axis refers to the days of admission relative to midnight on the first full day of admission. Units for each predictor are as follows: QRS duration (ms), platelet count (cells*10^−3^/uL), aPTT (seconds). Note that for all dynamic clinical data, values are assumed constant until a new measurement is made. *Abbreviations: white blood cell count (WBC), diastolic blood pressure (DBP), systolic blood pressure (SBP), glomerular filtration rate (GFR), activated partial thromboplastin time (aPTT)*

Supplementary Fig.S2 illustrates the COVID-HEART’s capability to accurately predict each CV outcome within outcome windows of different durations. This capability may provide significant clinical value in determining the patient’s short-term and longer-term risk, thus ensuring appropriate intervention and resources allocation. As the figures illustrate, validation and test results are comparable, indicating strong generalizability of the COVID-HEART. Fig.3 and Supplementary Fig.S3 provide examples of time-series clinical data and resulting risk scores for “true positive” and “true negative” predictions for patients in the test set for each CV outcome. Supplementary Fig.S4 illustrates two incorrect predictions; these are discussed in Supplementary Results.

In analyzing the results over many train-test splits, we found that for both outcomes, a larger number of sliding time windows in the test set were predicted positive for patients that eventually experienced the outcome as compared to those that did not: 43% (95% CI: 41%-45%) vs. 13% (95% CI: 12%-14%) for cardiac arrest, 58% (95% CI: 50%-65%) vs 27% (95% CI: 24%-30%) for thromboembolic events. This suggests that the model is sensitive in identifying warning signs of an impending adverse event earlier than the pre-specified outcome window (Supplementary Fig.S5). Indeed, the median early warning time for patients in the test set was 18 hours (IQR: 13-20 hours) for cardiac arrest and 72 hours (IQR: 12-204 hours) for thromboembolic events, although the classifier was trained to predict outcomes within 2 hours for cardiac arrest and 24 hours for thromboembolic events. This could represent a clinically useful “early warning” system.

It is essential for clinical decision-making to identify the features that most contribute to the predicted risk score for a particular CV outcome. We used a lasso regression algorithm to select features for prediction of cardiac arrest; this algorithm enforces sparsity and is appropriate for both continuous and categorical features. We used ANOVA F-value-based feature selection for prediction of thromboembolic event and strictly limited the number of features selected due to the small number of events in the development set. Table 1 lists the features with the largest coefficients in the optimal classifier for each of the two CV outcomes. Note that features were normalized prior to classifier training, and that models are not simple logistic regressions, thus interpretation of the coefficients is not straightforward. Many of these features confirm previous observations in cohorts of severely ill COVID-19 patients. For example, lower O_2_ saturation^15^ is associated with cardiac arrest and multiple coagulation-related labs results are associated with thromboembolic events.^27,28^ Interestingly, we found that the standard deviation of the QRS duration over the patient’s hospitalization up to the time of prediction was among the most influential in predicting thromboembolic events, which has not previously been reported; as of December 2020, most studies had only focused on the association of laboratory values with thromboembolic events.

The COVID-HEART risk prediction approach provides transparency and clinical explainability, including the ability to determine which features are dominant in a patient’s risk level at a particular time, which may suggest potential patient-specific targets for clinical intervention. Prediction models for CV adverse events in patients with COVID-19 have been limited by lack of sufficient data, impractical requirements for use (e.g. that all data be available for all patients or that measurements are taken at the same time relative to time of admission), and overly restrictive inclusion/exclusion criteria that result in an idealistic training cohort not representative of real patient data.^23,29^ Our model is designed to handle real-world data, which may include noise, missing variables, and data collected at different points in a patient’s hospitalization. The inclusion of multiple time-duration features gives the model the “memory” advantages of a long short-term memory neural network without compromising explainability or becoming a “black box”. It is trained in a manner that achieves high sensitivity *and* specificity despite severe class imbalance. To our knowledge, these techniques have not previously been combined in real-time predictors for CV events.

In this study we demonstrate highly accurate prediction of cardiac arrest and thromboembolic events in hospitalized COVID-19 patients using the continuously-updating COVID-HEART predictor. In its current implementation the predictor can facilitate practical, meaningful change in patient triage and the allocation of resources by providing real-time risk scores for CV complications occurring commonly in COVID-19 patients. The COVID-HEART can be re-trained to predict additional adverse CV events including myocardial infarction and arrhythmia. The potential utility of the predictor extends well beyond hospitalized COVID-19 patients, as COVID-HEART could be applied to the prediction of CV adverse events post-hospital discharge or used in pre-hospital emergency medical services. Additionally, the ML methodology utilized here could be expanded to use in other clinical scenarios that require screening or early detection, such as risk of hospital readmission, with the ultimate goal of improved clinical outcomes through early warnings and resultant opportunity for timely intervention.

## Data Availability

The authors declare that all data supporting the findings of this study are available within the paper and its supplementary information. The clinical data was all extracted from the JH-CROWN registry. Clinical data is available upon request at the following web address: https://ictr.johnshopkins.edu/coronavirus/jh-crown/. IRB approval is required to gain access to protected health information.

## Acknowledgments

The data utilized for this publication were from the Johns Hopkins COVID-19 Precision Medicine Center of Excellence, officially designated by the Precision Medicine Analytics Platform (PMAP) leadership, and based on the contribution of many patients and clinicians. This work would not be possible without the assistance of Dr. Paul Nagy, Bonnie Woods, Michael Cook, Brandon Smith, Philip Gianuzzi, Brian Garibaldi, and all other clinicians and data scientists at Johns Hopkins University who contributed to the COVID data resource design and collation. We are grateful to Dr. Vadim Zipunnikov for his helpful suggestions. This work was supported by a RAPID grant (2029603) from the National Science Foundation to N.A.T., a grant from the Lowenstein Foundation to N.A.T., and a National Science Foundation Graduate Research Fellowship (DGE-1746891) to J.S.

## Author Contributions

J.K.S, A.N.D, E.S, D.M.P. A.G.H, and N.A.T contributed to the study design. A.N.D, K.N.A, A.S.M, N.A.G and A.G.H provided clinical perspective and interpretation of the clinical variables. A.S.M, N.A.G, and A.G.H helped supervise and assisted in data collection. J.K.S ran experiments, recorded results, performed data analysis, created figures, and wrote the manuscript. N.A.T was the senior supervisor on all aspects of the project and contributed to the writing. All authors read, edited, and approved the submitted manuscript.

## Code Availability

The following open-source Python software libraries were used in data processing and analysis: numpy (version 1.18.1, available from: https://numpy.org/), pandas (version 1.1.4, available from: https://pandas.pydata.org/), imblearn (version 0.7.0, available from: https://imbalanced-learn.org/stable/), scipy (version 1.4.1, available from: https://www.scipy.org/), scikit-learn (version 0.23.2, available from: https://scikit-learn.org/stable/), and hcuppy (version 0.0.7, available from: https://pypi.org/project/hcuppy/). The code can be implemented using these packages with the methods detailed here.

## Methods

### Source of Data

To develop and validate the COVID-HEART predictor, we performed a retrospective study of patients in the JH-CROWN COVID-19 registry. Patient data was collected for this study from the registry between March 1, 2020 and September 27, 2020. Clinical data and outcomes were limited to those recorded in-hospital; there was no post-discharge follow-up. Patients were randomly assigned to development (80%) and testing (20%) data sets with stratification to ensure there were approximately the same proportion of patients with and without adverse CV events in each set. Assignment to development and testing sets was performed separately for prediction of cardiac arrest and thromboembolic events since inclusion/exclusion criteria and the proportion of patients with events were different. Supplementary Table S3 provides a demographic and clinical comparison of patients who did and did not experience each adverse event. Supplementary Table S4 provides a demographic and clinical comparison of patients in the development and testing set for each outcome.

### Patient Population

The JH-CROWN COVID-19 registry includes patients of all ages seen, since January 1, 2020, at any Johns Hopkins Medical Institution facility (inpatient, outpatient, in-person, video consult, or lab order) with confirmed COVID-19 or suspected of having COVID-19. The cohort is defined as having a completed laboratory test for COVID-19 (whether positive or negative), having an ICD-10 diagnosis of COVID-19 (recorded at the time of encounter, entered on the problem list, entered as medical history, or appearing as a billing diagnosis), or flagged as a “patient under investigation” for suspected or confirmed COVID-19 infection. Further details are available on the Johns Hopkins Institute for Clinical and Translational Research website.

Additional inclusion and exclusion criteria were applied for the COVID-HEART study, which resulted in a subset of the JH-CROWN registry being included. Supplementary Fig.S1 illustrates the flow of patients through the study. The COVID-HEART study included adult patients (age >=18 at the time of COVID-19 diagnosis) admitted as inpatients to any of the following hospitals in the Johns Hopkins Health System: Howard County General Hospital, Suburban Hospital, Sibley Memorial Hospital, Johns Hopkins Bayview Medical Center, and Johns Hopkins Hospital. For an admission to be included, patients must have had a laboratory-confirmed SARS-CoV-2 infection within 14 days prior to the date of admission or during the admission. The minimum length of time from admission to discharge or death was 4 hours for cardiac arrest prediction and 72 hours for prediction of thromboembolic events, the difference being necessitated by the time granularity with which each outcome could be identified, discussed in further detail in the following section. Time spent in the emergency department did not count towards the admission duration, but if a patient had clinical data (e.g. laboratory values or vital signs) recorded in the emergency department prior to admission, those values were used to initialize the clinical data inputs at the start of their inpatient admission. Data were censored at the time of outcome or discharge.

Multiple admissions were handled as follows. If a patient was transferred between hospitals in the health system and thus had two admissions recorded in the JH-CROWN registry with a gap of fewer than 4 hours, it was treated as a single admission. However, if a patient was discharged and re-admitted to the same hospital or a different hospital more than 4 hours later, the admissions were treated separately, and all dynamic clinical data inputs were “reset” for the second admission. Admission-based inclusion/exclusion criteria were applied separately for each admission.

Additional exclusion criteria were applied for prediction of thromboembolic events. Patients were excluded if they experienced a thromboembolic event immediately prior to admission that was diagnosed on admission or within 24 hours of admission. For prediction of both outcomes, patients were not excluded based on treatments received, disease severity, need for intensive care, missing clinical variables, or any other reason not listed here. Although excluding patients for these reasons may have improved the models’ performance, it would have resulted in an unrealistically “clean” cohort not representative of real clinical data, making the risk predictor less useful in a real-world clinical setting.

### Outcome Definition

In-hospital cardiac arrest was defined according to the time of death recorded in the JH-CROWN database. 270 cardiac arrests were associated with mortality and were defined by querying the time of death, however, cardiac arrests with resuscitation were also included in the outcome definition. These were queried by searching for the ICD-10 code ‘I46.X’ within the problem list and encounter diagnosis list. Eight patients had ICD-10 diagnosis codes reflecting in-hospital cardiac arrest with resuscitation. Overall, 277 out of 2178 patients (12.7%) experienced cardiac arrests; one patient experienced a cardiac arrest with resuscitation and later died. For patients with multiple cardiac arrests, the first outcome was used, and the remainder of their data were censored.

Thromboembolic outcomes included pulmonary embolism confirmed on computed tomography (CT) angiography of the chest, non-hemorrhagic stroke confirmed on CT of the head, and deep venous thrombosis confirmed on either vascular ultrasound or CT of the abdomen or pelvis. Findings that were diagnosed or clinically apparent on initial presentation (confirmed on imaging within 24 hours of presentation) were excluded from analysis. For a patient with multiple adverse coagulation outcomes during their hospitalization, the first outcome was used. We note that such a strict outcome definition could mean that some outcomes were missed, especially if a patient’s immediate cause of death was a thromboembolic event or if the event was confirmed by point-of-care ultrasound that was not recorded in the imaging procedure list. However, we found that alternative outcome definition methods (such as ICD-10 diagnosis codes) resulted in many “false positive” outcomes upon chart review, so this method was chosen to ensure all thromboembolic events were confirmed with a consistent, objective level of clinical certainty. Overall, 32 out of 1601 (2.0%) eligible patients experienced imaging-confirmed thromboembolic events. 16 additional patients (1.0%) had imaging-confirmed thromboembolic events recorded on admission or within 24 hours of admission and were excluded for that reason.

### Predictors

Supplementary Table S2 lists all clinical data inputs from which predictors were extracted. Here, we discuss the definition of these predictors, how they were measured, and pre-processing steps undertaken prior to dynamic feature extraction.

Demographic inputs included age, gender, weight, height, body mass index, and race. Gender was defined as the patient’s legal gender (Male or Female) as listed in the electronic health record (EHR). Race was self-reported and divided into three categories according to the most common values in the JH-CROWN registry: Black, white, and other. The inclusion of race in machine learning models is controversial.^32^ However, there is significant evidence that Black patients and other patients of color experience worse outcomes in COVID-19.^33^ We were concerned that by not including race, our model may fail to account for a higher baseline risk of adverse outcomes among Black patients in the study cohort’s geographic area.^34^ Future work, prior to a prospective study, could include a re-analysis of the current results to ensure that the predictions are not systematically less accurate for any demographic group.^32^ Comorbidities were defined by mapping ICD-10 codes according to the Elixhauser comorbidity definitions^31^ using the hcuppy python library.^35^

Vital signs were extracted from flowsheet data recorded in the EHR and added to the JH-CROWN registry. Pulse measurements were excluded if the recording was 0. Both systolic blood pressure (SBP) and diastolic blood pressure (DBP) were recorded using either a blood pressure cuff or an arterial line. These were combined into a single input. If a given time point had measurements for SBP and DBP with both modalities, the arterial line measurement took priority. SBP measurements between 30 and 270 mmHg were considered valid. DBP measurements between 30 and 130 mmHg were considered valid. If the difference between SBP and DBP was less than 15 mmHg, both measurements were considered invalid. Respiratory rates between 4 and 52 breaths per minute were considered valid. Temperatures between 89°F and 105°F (31.7°C – 40.6°C) were considered valid. Pulse oxygen saturation between 30% and 100% was considered valid. Other flowsheet data, such as fraction of inspired oxygen and positive end expiratory pressure, were not included as these are directly influenced by a physician’s assessment of the patient’s condition, rather than physiologic data reflecting the patient’s condition in an unbiased manner. Heart rhythm indicators were also extracted from flowsheet data.

Laboratory tests results were extracted from EHR data and were time-stamped at the time the result was received, not the time of collection. This was done to ensure the model was trained with realistic data; in a prospective study it would not be possible to know the result of a laboratory test for a patient at the time the specimen would be collected.

ECG measurements were extracted from the 12-lead ECG. As with laboratory tests, these measurements were time-stamped at the time the result was received, not the time of the procedure. Parameters (QRS duration, QT interval, etc.) were evaluated by the clinician who interpreted the ECG results.

For all clinical data inputs, outliers that were >5 standard deviations from the mean were removed. This threshold was chosen to avoid excluding abnormal but non-erroneous values. We intentionally applied minimal “corrections” to clinical data inputs to ensure our development and validation data sets were realistic and that our model could be applied in a real-world clinical setting.

The testing data set was identified and sequestered from the training data prior to model development. Since this was a retrospective study and did not include any data collected prospectively, there was no need of blind assessment of predictors for patients in the testing set. Patients were assigned to training and test sets after predictors were collected and outcomes were defined, but prior to model development.

### Sample Size

The study size was determined by the number of patients in the JH-CROWN registry who met all inclusion and exclusion criteria for prediction of each outcome. 20% of the data was held out for testing; this was pre-determined.

### Feature Extraction and Missing Data

Here we present methods for extracting features from dynamic clinical data and handling of missing predictors in the analysis. All pre-processing steps were performed using the Python *Pandas* data analysis library. Laboratory tests, vital signs, and ECG measurements were handled similarly. For each patient, each measurement for each variable within these categories was associated with a time-stamp at which the measurement was received. Data were re-sampled in 30-minute increments for the prediction of cardiac arrest and in 1-hour increments for the prediction of thromboembolic events with mean interpolation if multiple measurements were made in a given window. Missing values from the beginning of the patient’s hospitalization (e.g., if they did not have a measurement for a particular laboratory test until hour 48, or at any point during their hospitalization) were left empty and handled later, within the modeling pipeline. Missing values following a measurement (e.g., if a patient had an ECG at hour 12, then did not have another ECG until hour 48) were handled with forward filling; each variable was held constant until a new measurement was made.

In the remainder of the Methods, we refer to “time point”, “time window”, “feature window”, “outcome window”, and “positive” time window. A time point indicates a single moment in time. The time window before a time point, during which clinical data are collected and features are extracted, is referred to as the “feature window”. The time window immediately after, in which the risk of a particular CV outcome is predicted, is referred to as the “outcome window”. “Positive time windows” or “positive time points” are time windows or points for which the patient experienced the CV outcome of interest in the following outcome window.

Following the preprocessing steps described above, dynamic features were calculated from the processed time-series clinical data inputs as illustrated in Fig.1B. “Short features” encompassed a short window of time immediately preceding the time point at which the prediction was to be made. For example, if the feature window length was 2 hours, these features would include the mean, standard deviation, minimum, maximum, and amplitude of first frequency in Fourier space of the variable over the preceding 2 hours. “Long features” included the mean, standard deviation, minimum, and maximum over the patient’s entire hospitalization preceding the time point at which the prediction was to be made. “Exponentially weighted decay features” also encompassed the patient’s entire hospitalization preceding the time point at which the prediction was to be made, but the measurements were exponentially weighted according to how recently they were made with more recent measurements weighted more strongly and a half-life of 1 day.

Heart rhythm indicators were re-sampled similarly to other dynamic clinical data inputs but were treated discretely. For each window, two variables were recorded for each heart rhythm indicator (Atrial fibrillation, heart block, etc.): a binary indicator of whether the patient experienced that heart rhythm within the window and an integer-valued variable indicating how many times that heart rhythm was noted within the window. It was assumed that if a patient did not have any heart rhythm annotations within a particular hour, they did not experience an abnormal heart rhythm during that window, so missing values were filled in with zero for both the binary indicator variable and integer-valued variable. “Short features” and “long features” were calculated for each heart rhythm indicator but included only the sum (total number of times each was recorded over the interval) and maximum (maximum number of times a rhythm was recorded in a single hour within the interval).

Dynamic features were extracted at each time point during each patient’s hospitalization. The time-step between time points at which predictions were made was 1 hour for prediction of cardiac arrest and 24 hours for prediction of thromboembolic events. For thromboembolic events, each time window began at midnight; for cardiac arrest, each time window began at the top of the hour, commencing with the first full hour after the patient was admitted as an inpatient. The difference in time-step was due to the difference in the time granularity of the outcome labels. Although cardiac arrest outcomes could be defined by the minute in which they occurred, and thus it would be appropriate to use a time-step as small as 1 minute, 1 hour was chosen to balance computational costs with the desire to train the classifier with as much data as possible. A time-step of 1 hour resulted in 412,198 time windows for the development set, which produced an accurate, generalizable classifier as demonstrated by the strong cross-validation and testing results for prediction of cardiac arrest.

Each time-point was labeled with a binary outcome label indicating whether the patient experienced the outcome of interest in an “outcome window” following the time-point. 24 hours was selected as the outcome window for prediction of thromboembolic events as this was the minimum interval in which outcomes could be identified. 2 hours was selected as the outcome window for prediction of cardiac arrest based on practical clinical considerations. A “2-hour warning” for impending cardiac arrest would provide healthcare personnel sufficient time for intervention if indicated. Static features, including demographics and comorbidities, were then concatenated to the dynamic features. These were constant for all time points for each patient.

### Statistical Analysis Methods

Three linear classifier configurations were investigated for prediction of each outcome using the feature windows and outcome windows described above (both 2-hour windows for cardiac arrest and 24-hour windows for thromboembolic events): one with short features only, one with all feature types, and one with all feature types and an additional pre-processing step to re-sample the training data to handle class imbalance. Here, we discuss the specifications for each model. Unless otherwise stated, methods were the same for all three classifier configurations.

#### Predictor Handling During Development

After extracting features for all patients in the development set, static and dynamic features which were missing for >60% of time windows were removed. Pre-processing steps were integrated into a scikit-learn *Pipeline* to ensure data were handled correctly according to whether they were in the training or validation set during cross-validation and testing. For all three classifier configurations, the pre-processing steps included mean-value imputation for numerical features that were missing (typically at the beginning of a patient’s hospitalization or if a certain laboratory test was never performed for a given patient), scaling all numerical features to zero mean and unit variance, and feature selection using a lasso regression model for sparsity. This feature selection method was chosen as it is not biased towards selecting high-cardinality variables over variables with fewer discrete values (e.g., binary comorbidity features), in contrast with other popular feature selection methods such as the random forest algorithm. We used ANOVA F-value-based feature selection for prediction of thromboembolic events and significantly restricted the number of features that could be selected to reduce the likelihood of over-fitting due to the very small number of events in the development set.

For the classifier configuration with re-sampling, two additional steps were added to the pipeline after scaling but before feature selection. First, the Synthetic Minority Oversampling TEchnique (SMOTE) for nominal and continuous variables^25^ was applied to up-sample the positive time windows by synthesizing new examples from the existing positive time windows. This was followed by random under-sampling to down-sample the negative time windows. There were typically only about 0.1-1.0% positive time windows prior to re-sampling, depending on the outcome window and outcome type, so this step was applied to improve the class imbalance during training.

#### Model Specification

The model evaluated was a linear classifier trained with stochastic gradient descent. This model was chosen as it is highly explainable (not a “black box”), it is efficient to train with hundreds of thousands of time windows, and it can be updated without requiring full re-training. As COVID-19 treatment paradigms change, we expect that model updating would be necessary to retain accuracy among evolving clinical practices. The learning rate of the model was set to “optimal” with early stopping and balanced class weight. The model was the final step in the scikit-learn *Pipeline*.

Five-fold stratified group cross-validation was used to optimize hyperparameters of the COVID-HEART predictor. Groups were set such that all time points from each patient were held out in the same fold of cross-validation. Hyperparameters were optimized for all steps in the pipeline with 500 iterations of a random grid search for prediction of thromboembolism (since the time step was 24 hours, there were fewer time windows and thus training was more efficient, especially for the model with re-sampling) and 100 iterations for prediction of cardiac arrest to maximize the validation AUROC. Hyperparameters included the over- and under-sampling strategy for the model with re-sampling, the maximum number of features selected, the loss function of the linear classifier (hinge, log, modified Huber, Huber, squared hinge), the regularization penalty (L1, L2, and L1L2), the regularization strength, and the L1 ratio for L1L2 regularization. Losses were weighted during training to strongly penalize errors for positive time windows. Following training, the optimized classifier was calibrated to provide risk probabilities in addition to binary predictions.

#### Model Testing

Following design of feature extraction methods, model development, and model training, the optimal models for prediction of each outcome were re-fit and calibrated using the entire development set. Static and dynamic features were then calculated for patients in the testing set using the same methods as for the development set. The fitted models were used to predict the risk of each CV outcome at each time point for each patient in the testing set. A binary prediction was also made at each time point using the optimal threshold determined by the development data during training.

Although predictions were made at the same time steps for patients in the test set for consistency with the development set, it is possible to apply the model at any arbitrary time during a patient’s hospitalization. We envision that in practice, it could provide the physician with an updated risk score each time any new clinical data input becomes available or only after passing a certain “high risk” threshold, to reduce healthcare provider “alert fatigue”.

#### Model Performance Assessment

Model performance was assessed by the following metrics: accuracy, balanced accuracy, sensitivity, specificity, AUROC, F1-score, and precision. These were calculated over the folds of cross-validation. Models were optimized and compared based on the cross-validation AUROC. The output of the classifier was a risk score (predicted probability that the patient will experience a particular CV event in the outcome window). A binary risk threshold was selected automatically to balance sensitivity and specificity for the development data, then applied to the test data to generate binary predictions for each time window in the test set. These binary predictions were used to calculate the classification performance metrics. This was repeated for each fold of cross-validation and for the entire development set to make predictions for the separate test set. Additional metrics were calculated based on the binary risk determination at each time point to further investigate the sensitivity of the model in identifying early warning signs of an impending adverse CV event. These included the mean, median, and standard deviation of early warning time, defined as the number of hours prior to an event for which the binary risk prediction was positive. As a secondary analysis, the number of time windows predicted positive for patients who eventually experienced events and for patients who did not were compared.

#### Model Updating

The testing set was sequestered until the end of model development. There were no changes made to the model following testing. After determining the optimal classifier configuration for prediction of each event within the outcome windows specified above, we performed a secondary analysis in which we varied the length of the outcome window to investigate whether the COVID-HEART predictor could forecast outcomes within multiple intervals. At this point, the feature extraction and modeling methodology was pre-determined and only the outcome window were varied.

#### Nested Cross-Validation

Following training and validation of the optimal classifier configuration for prediction of each outcome with the train-test split discussed above, we added an outer loop of cross-validation to repeat the training and testing many times. Each full iteration of cross-validation included 5 train-test splits, selected randomly without replacement, to avoid bias. This was done to calculate the mean and 95% confidence interval for each validation and testing performance metric. Since there were few events for each outcome, the original train-test split results may be a poor reflection of the COVID-HEART predictor’s true capabilities. Repeating the train-test split provided a more accurate estimate of the models’ cross-validation and test performance. All test patient example predictions and data describing the characteristics of the development and testing sets were generated using the “original” optimal model with the initial randomly selected development and testing sets.

### Development vs. Validation

There were no differences between development and test data in setting, eligibility criteria, outcome, and predictors. All patients were from a subset of the JH-CROWN registry and were randomly assigned to development and test sets *after* predictors and outcomes had been collected.

### Limitations

While the COVID-HEART predictor overcomes many limitations of previously developed models for risk assessment in COVID-19 and is trained and tested on a large cohort of patients from multiple hospitals, several limitations remain. These include the lack of prospective validation in an external cohort and the requirement for imaging confirmation of thromboembolic events. The latter means that some thromboembolic events may not have been recorded, including those that were subclinical or were the immediate cause of death.

There are also inherent limitations in the use of registry data rather than data prospectively collected for the purposes of this study.^36^ These include the potential for measurement error, inaccurate patient-reported history (e.g. smoking), and missing data. There is also left censoring measurement bias (e.g. patients transferred from other health systems may have different types of data available, since only data collected within the Johns Hopkins health system is available in the registry) and, since Johns Hopkins is a tertiary care center, patients in the registry tend to have a more severe disease course than the general population. However, the COVID-HEART predictor, designed to handle real-world clinical data, achieved strong results despite these limitations.

Additional limitations stem from the use of the JH-CROWN registry and were not easily overcome. These include confounding by indication, which means that treatments were selected based on clinical indication. While our model did not include treatments or other variables that were directly influenced by clinical indication, some variables in the model were likely indirectly influenced by clinical indication. For example, the pulse oxygen saturation may have been affected by changes in ventilator settings for patients who were receiving mechanical ventilation. There is also a subgroup of patients who had pre-existing DNR/DNI/comfort care orders. These patients would have received no interventions leading up to an adverse CV event, which means that the sequalae of physiologic changes for these patients may be different than for patients who received interventions prior to an adverse CV event. Finally, there is selection bias inherent to including only patients who sought care at a hospital; patients without insurance, undocumented patients, and patients with other barriers to seeking care may be less likely to be included.

## Supplementary Results

### Participants

Supplementary Fig.S1 shows the flow of participants through the study. 2178 patients met eligibility criteria for cardiac arrest prediction. 277 experienced cardiac arrest. Supplementary Table S3 provides a clinical and demographic comparison of patients who did and did not experience cardiac arrest. Overall, patients who experienced cardiac arrest were older (mean age 73.7 years vs. 59.5 years, p<0.001) and spent more time in the hospital (338.8 hours vs. 229.2 hours, p<0.001). They were more likely to have valvular disease (9.7% vs. 5.1%, p=0.003), peripheral vascular disorders (14.1% vs. 8.2%, p=0.002), neurological disorders (39.0% vs. 18.5%, p<0.001), iron deficiency anemia (30.3% vs. 21.8%, p=0.002), hypertension without complications (70.0% vs. 55.4%, p<0.001), congestive heart failure (27.1% vs. 14.3%, p<0.001), fluid and electrolyte disorder (37.9% vs. 24.1%, p<0.001), and a history of smoking (20.2% vs. 14.6%, p=0.019). In investigating their first laboratory measurements on admission to the hospital for a select subset of laboratory tests that have been shown to be associated with adverse outcomes in COVID-19, patients who experienced cardiac arrest had statistically significantly higher NT-pro-brain natriuretic peptide (pro-BNP), white blood cell count, D-Dimer, C-reactive protein, ferritin, and troponin. They had statistically significantly lower absolute lymphocyte count. Of note, many patients were missing measurements for several of these tests. Finally, in investigating their first ECG recordings on admission to the hospital, patients who experienced cardiac arrest had statistically significantly higher QTc interval, T axis, ventricular rate, and atrial rate.

These patients were divided with stratified random selection into a development set of 1742 patients (80%) in which 222 (12.7%) experienced cardiac arrest and a testing set of 436 patients (20%) in which 55 (12.6%) experienced cardiac arrest. Supplementary Table S4 provides a comparison between the randomly selected development and testing sets. Since these patients were randomly selected, there were a few spurious differences between the development and validation set but no systematic differences in demographic or clinical characteristics.

1617 patients met eligibility criteria for thromboembolic event prediction. 16 of these patients were excluded for having a thromboembolic event within 24 hours of admission, this usually indicated that the event occurred prior to admission and was confirmed with imaging on admission. 32 of the remaining 1601 experienced imaging-confirmed in-hospital thromboembolic events. Supplementary Table S3 provides a clinical and demographic comparison of patients who did and did not experience thromboembolic events. Patients who experienced thromboembolic events had longer admission duration (933.0 hours vs. 297.0 hours, p<0.001). They were more likely to have pulmonary circulation disorders (40.6% vs. 5.3%, p<0.001) and congestive heart failure (34.4% vs. 18.4%, p=0.039). On admission, they had statistically significantly lower absolute lymphocyte count and statistically significantly shorter QRS duration.

These patients were divided with stratified random selection into a development set of 1280 patients (80%) in which 26 (2.0%) experienced imaging-confirmed thromboembolic events and a testing set of 321 patients (20%) in which 6 (1.9%) experienced imaging-confirmed thromboembolic events. Supplementary Table S4 provides a comparison between the randomly selected development and testing sets. There were no statistically significant differences in any clinical or demographic characteristics between the development and testing sets.

Supplementary Table S3 indicates the number of patients for which each measurement was missing. This does not necessarily mean they never had a measurement for a certain variable. It may mean that they had a recording at a hospital in a different health system prior to being transferred to a hospital in the Johns Hopkins Health System or that data was missing from the JH-CROWN registry. This is an inherent limitation in the use of retrospective registry data, discussed in further detail in Methods.

### Model Specification

The optimal model for prediction of cardiac arrest with a feature window of 2 hours, outcome window of 2 hours, and time step of 1 hour was a classifier with short features only. The optimal hyperparameters included a maximum of 31 features selected, log loss, L2 regularization penalty, and regularization strength of 0.10. The optimal model for prediction of thromboembolic outcomes with a feature window of 24 hours, outcome window of 24 hours, and time step of 24 hours was a classifier with all features and no re-sampling. The optimal hyperparameters included 3 features, Huber loss, L1L2 regularization penalty, L1 ratio of 0.95, and regularization strength of 0.01.

Table 1 lists the features with largest absolute coefficients in the model for prediction of each outcome along with their values for time windows in the development and test sets. Feature selection was performed using the development set only. The most important features for prediction of cardiac arrest within 2 hours included chronic lung disease, many vital signs, lab tests that indicate inflammation and metabolic function, and heart rhythm annotations that provide information about cardiac function. Several of these have previously been noted as predictors of various adverse outcomes in COVID-19.^21,26,37,38^ For example, elevated fibrinogen has been linked to high plasma viscosity in COVID-19, which may contribute to morbidity and mortality.^28^ This serves as a “sanity check” that the model is learning reasonable associations between predictors and outcomes, despite its novel real-time nature. The optimal classifier configuration included dynamic features from the 2 hours prior to the time point of prediction, suggesting that the decline in function leading to cardiac arrest occurs rapidly.

The features with largest absolute coefficients for prediction of thromboembolic events within 24 hours were derived from QRS duration, platelet count, and activated partial thromboplastin time (aPTT). Other variables, including D-Dimer, IL-6, and Troponin I were associated with thromboembolic events (Supplementary Table S1), but only a few features could be included in the model due to the small number of events in the development set. One of the three features included in the model is a short feature calculated using clinical data from the 24 hours prior to the time of prediction and two are long features encompassing the patient’s entire hospitalization prior to the time of prediction. This suggests that data from only the day prior to the time of prediction do not tell the full story of the patient’s risk.

APTT is associated with use of anti-coagulation therapy, suggesting that the model may be implicitly learning information about the physician’s assessment of the patient’s condition. Interestingly, to our knowledge, as of December 2020, there have been no studies describing the association of ECG abnormalities with thromboembolic outcomes in COVID-19. In our study, the standard deviation of the patient’s QRS duration over their entire hospitalization up to the time of prediction was among the three selected for inclusion in the classifier for prediction of thromboembolic events, however it is unclear whether these abnormalities were related to cardiac stunning, medications, or other etiologies. The results of our study suggest possible avenues for future research.

### Model Performance

The overall performance of the optimal model for prediction of each outcome is discussed in the main text results. Here, we discuss the results in more detail, including patient-specific example predictions for patients in the test set for each outcome.

#### Test Patient Example Predictions

The first example is the “true positive” prediction for one patient in the test set for each outcome, as shown in Fig.3. In predicting the risk of cardiac arrest within 2 hours for the patient whose data is shown in Fig.3A, results show that their risk is very low for the first 11 days of their hospitalization. On the 11^th^ day, their oxygen saturation drops rapidly, and their systolic blood pressure decreases, while their white blood cell count increases. The effects of these changes are reflected in the patient’s risk score, which increases very rapidly in the 2 hours leading up to the time at which the patient experienced cardiac arrest. Although there are isolated changes in the risk score inputs during the first 11 days of their hospitalization, the COVID-HEART predictor is successful in determining when the patient becomes at risk of impending cardiac arrest by considering all the inputs together.

In predicting the risk of a thromboembolic event within 1 day for the patient whose data is shown in Fig.3B, the results demonstrate that the risk score is low for the first 4 days of the patient’s hospitalization, then it crosses the binary risk threshold, continues to rise until day 7, and remains steadily above the binary risk threshold until day 14, when the patient experienced an imaging-confirmed thromboembolic event. The patient has steadily increasing platelet count, which was found to be predictive of thromboembolic events in our study, though to our knowledge it has not been reported elsewhere. The patient also has elevated aPTT at multiple points during their hospitalization, which may suggest that the physician treated the patient with heparin. The risk predictor can identify that the patient is at risk for an event by considering the changes in features extracted from these inputs over the entire duration of the patient’s hospitalization as well as in the day leading up to the event. This highlights the usefulness of the COVID-HEART risk predictor in identifying at-risk patients that may not have raised clinical suspicion for an impending thromboembolic event based on traditional risk markers.

Supplementary Fig.S3 shows an example of a “true negative” prediction for one patient in the test set for each outcome. The cardiac arrest risk score for the patient whose data is shown in Supplementary Fig.S3A remains below 0.05% for their entire hospitalization. This patient has several drops in both systolic and diastolic blood pressure, but the COVID-HEART risk predictor successfully assesses their risk as low. This also illustrates the COVID-HEART risk predictor’s ability to cope with missing clinical data; the patient has no measurements for fibrinogen during their hospitalization. This patient did not experience cardiac arrest and was discharged after 2 days in the hospital. The thromboembolic event risk score for the patient whose data is shown in Supplementary Fig.S3B remains below 0.2% for their entire hospitalization. This patient did not experience any imaging-confirmed thromboembolic events.

Supplementary Fig.S4 shows an example of an incorrect prediction for one patient in the test set of each outcome. The patient whose clinical data is shown in Supplementary Fig.S4A experienced cardiac arrest on day 11 of their hospitalization. Their risk score for cardiac arrest increased rapidly on day 8 corresponding to a sharp decrease in their systolic and diastolic blood pressures. However, it then decreased and remained at a constant, slightly elevated level for the following 3 days prior to the time at which they experienced cardiac arrest. Although they were not at the highest risk immediately before they experienced cardiac arrest, their risk score was above the threshold for cardiac arrest risk determined by the development data at the time they experienced cardiac arrest predictor. Thus, although this was technically a correct prediction, we focus on the risk score spike on day 8 as an example of a false positive prediction.

The patient whose clinical data is shown in Supplementary Fig.S4B experienced an imaging-confirmed thromboembolic event on day 4 of their hospitalization. Their risk score was low for the duration of their hospitalization. It is unknown why the COVID-HEART predictor was unsuccessful for this patient. Although the COVID-HEART predictor is trained to cope with missing data, it is possible that the predictor was less accurate in this case because the patient had few measurements for QRS duration and platelet count. The patient’s aPTT was elevated above normal levels, which may indicate the patient was receiving prophylactic heparin. This highlights the need for further investigation of incorrect predictions by the COVID-HEART predictor.

#### Predicting CV Events Within Various Outcome Windows

After determining the optimal classifier configuration for prediction of each outcome with pre-determined outcome windows and short feature windows, we performed a series of experiments in which we varied the duration of the outcome window and repeated the training, optimization, and validation process as described in Methods. Supplementary Fig.S2 shows the results of varying the outcome window for prediction of each outcome. For prediction of cardiac arrest, the outcome window can vary from 1 to 24 hours with little change in AUROC, sensitivity, and specificity. This analysis shows that the COVID-HEART predictor can predict cardiac arrest within multiple outcome window durations, representing a continuous early warning system for cardiac arrest that may be able to determine both the patient’s short-term and longer-term risk.

Supplementary Fig.S2 presents numerical results for all outcome windows for the prediction of thromboembolic events. When the feature window is held constant at 24 hours, we see that the results are similar for prediction of thromboembolic events within 1, 2, 3, and 4 days. There were only 26 patients in the development set with imaging-confirmed thromboembolic events and these outcomes could only be identified per-day, not at the exact time they occurred, as with cardiac arrest. As a result, only a few features could be selected; it is possible that a larger feature set would lead to more accurate prediction of the patients’ risk of thromboembolic events since more details of the patients’ clinical states could be considered. We believe that with either a larger cohort (with more imaging-confirmed thromboembolic events) or re-assessing the outcome definition to include events confirmed by clinical suspicion rather than strictly requiring imaging confirmation could improve the results to a level comparable to the results for prediction of cardiac arrest. However, despite these limitations, the COVID-HEART predictor can accurately forecast thromboembolic events with a cross-validation AUROC of 0.77 (95% CI: 0.75-0.79) and a testing AUROC of 0.70 (95% CI: 0.67-0.73).

## Supplementary Figures/Tables

**Supplementary Table S1.** TRIPOD Checklist for development and validation of a multi-variable prognostic model.

| Section/Topic |  |  | Checklist Item | Page |
| --- | --- | --- | --- | --- |
| Title and abstract |  |  |  |  |
| Title | 1 | D;V | Identify the study as developing and/or validating a multivariable prediction model, the target population, and the outcome to be predicted. | 1 |
| Abstract | 2 | D;V | Provide a summary of objectives, study design, setting, participants, sample size, predictors, outcome, statistical analysis, results, and conclusions. | 2 |
| Introduction |  |  |  |  |
| Background and objectives | 3a | D;V | Explain the medical context (including whether diagnostic or prognostic) and rationale for developing or validating the multivariable prediction model, including references to existing models. | 2 |
|  | 3b | D;V | Specify the objectives, including whether the study describes the development or validation of the model or both. | 2, 3 |
| Methods |  |  |  |  |
| Source of data | 4a | D;V | Describe the study design or source of data (e.g., randomized trial, cohort, or registry data), separately for the development and validation data sets, if applicable. | 3, 12 |
|  | 4b | D;V | Specify the key study dates, including start of accrual; end of accrual; and, if applicable, end of follow-up. | 3, 12 |
| Participants | 5a | D;V | Specify key elements of the study setting (e.g., primary care, secondary care, general population) including number and location of centres. | 3, 12 |
|  | 5b | D;V | Describe eligibility criteria for participants. | 3, 12 |
|  | 5c | D;V | Give details of treatments received, if relevant. | N/A |
| Outcome | 6a | D;V | Clearly define the outcome that is predicted by the prediction model, including how and when assessed. | 13 |
|  | 6b | D;V | Report any actions to blind assessment of the outcome to be predicted. | N/A |
| Predictors | 7a | D;V | Clearly define all predictors used in developing or validating the multivariable prediction model, including how and when they were measured. | 13-14,24 |
|  | 7b | D;V | Report any actions to blind assessment of predictors for the outcome and other predictors. | 14 |
| Sample size | 8 | D;V | Explain how the study size was arrived at. | 14 |
| Missing data | 9 | D;V | Describe how missing data were handled (e.g., complete-case analysis, single imputation, multiple imputation) with details of any imputation method. | 14-15 |
| Statistical analysis methods | 10a | D | Describe how predictors were handled in the analyses. | 15 |
|  | 10b | D | Specify type of model, all model-building procedures (including any predictor selection), and method for internal validation. | 3, 15-16 |
|  | 10c | V | For validation, describe how the predictions were calculated. | 16 |
|  | 10d | D;V | Specify all measures used to assess model performance and, if relevant, to compare multiple models. | 16-17 |
|  | 10e | V | Describe any model updating (e.g., recalibration) arising from the validation, if done. | 17 |
| Risk groups | 11 | D;V | Provide details on how risk groups were created, if done. | N/A |
| Development vs. validation | 12 | V | For validation, identify any differences from the development data in setting, eligibility criteria, outcome, and predictors. | 17 |
| Results |  |  |  |  |
| Participants | 13a | D;V | Describe the flow of participants through the study, including the number of participants with and without the outcome and, if applicable, a summary of the follow-up time. A diagram may be helpful. | 3, 29 |
|  | 13b | D;V | Describe the characteristics of the participants (basic demographics, clinical features, available predictors), including the number of participants with missing data for predictors and outcome. | 18, 25-26 |
|  | 13c | V | For validation, show a comparison with the development data of the distribution of important variables (demographics, predictors and outcome). | 27-28 |
| Model development | 14a | D | Specify the number of participants and outcome events in each analysis. | 3, 13 |
|  | 14b | D | If done, report the unadjusted association between each candidate predictor and outcome. | N/A |
| Model specification | 15a | D | Present the full prediction model to allow predictions for individuals (i.e., all regression coefficients, and model intercept or baseline survival at a given time point). | 10 |
|  | 15b | D | Explain how to the use the prediction model. | 16-17 |
| Model performance | 16 | D;V | Report performance measures (with CIs) for the prediction model. | 8 |
| Model-updating | 17 | V | If done, report the results from any model updating (i.e., model specification, model performance). | N/A |
| Discussion |  |  |  |  |
| Limitations | 18 | D;V | Discuss any limitations of the study (such as nonrepresentative sample, few events per predictor, missing data). | 17 |
| Interpretation | 19a | V | For validation, discuss the results with reference to performance in the development data, and any other validation data. | 19-20 |
|  | 19b | D;V | Give an overall interpretation of the results, considering objectives, limitations, results from similar studies, and other relevant evidence. | 3-4, 19-20 |
| Implications | 20 | D;V | Discuss the potential clinical use of the model and implications for future research. | 3-4 |
| Other information |  |  |  |  |
| Supplementary information | 21 | D;V | Provide information about the availability of supplementary resources, such as study protocol, Web calculator, and data sets. | 12-33 |
| Funding | 22 | D;V | Give the source of funding and the role of the funders for the present study. | 4 |

**Supplementary Table S2:** Complete list of clinical data from which features were derived. These are discussed in further detail in Methods. Comorbidities are defined using ICD-10 codes according to the Elixhauser comorbidity definitions.^31^

| Category | Clinical Data Inputs |
| --- | --- |
| Demographics (6) | Age, gender, weight, height, body mass index, race |
| Comorbidities (30) | Current smoker, history of smoking, chronic pulmonary disease, diabetes mellitus with complications, diabetes mellitus without complications, lymphoma, valvular disease, psychosis, peripheral vascular disorder, pulmonary circulation disorders, hypothyroidism, alcohol abuse, neurological disorders, deficiency anemia, renal failure, liver disease, rheumatoid arthritis/collagen, solid tumor without metastasis, metastatic cancer, drug abuse, depression, HIV/AIDS, hypertension with complications, hypertension without complications, obesity, coagulopathy, peptic ulcer disease, congestive heart failure, paralysis, fluid and electrolyte disorders |
| Vital signs (6) | Pulse, systolic blood pressure, diastolic blood pressure, respiratory rate, temperature, pulse oxygen saturation |
| Lab tests (39) | NT-pro-brain natriuretic peptide (ProBNP), white blood cell count, absolute lymphocyte count, D-dimer, lymphocytes %, heparin (anti-Xa), interleukin-6 (IL-6) serum, C-reactive protein, ferritin, fibrinogen, troponin I, troponin T, bands, creatinine serum, creatinine plasma, blood urea nitrogen (BUN), BUN/creatinine ratio, sodium, potassium, hematocrit, glomerular filtration rate, glucose, chloride, carbon dioxide, calcium, procalcitonin, magnesium, alanine aminotransferase, aspartate aminotransferase, bilirubin, lactate whole blood, erythrocyte sedimentation rate, lactate dehydrogenase, platelet count, mean platelet volume, activated partial thromboplastin time, prothrombin time, anion gap, mean corpuscular volume |
| ECG measurements (9) | PR interval, QRS duration, QT interval, QTc interval, P axis, QRS axis, T axis, ventricular rate, atrial rate |
| Heart rhythm indicators (16) | Atrial fibrillation, atrial flutter, sinus arrhythmia, ectopy, heart block, sinus tachycardia, sinus bradycardia, junctional rhythm, ventricular tachycardia, asystole, ventricular paced rhythm, normal sinus rhythm, atrial paced rhythm, A-V sequential paced rhythm, supraventricular tachycardia, agonal |

**Supplementary Table S3:**
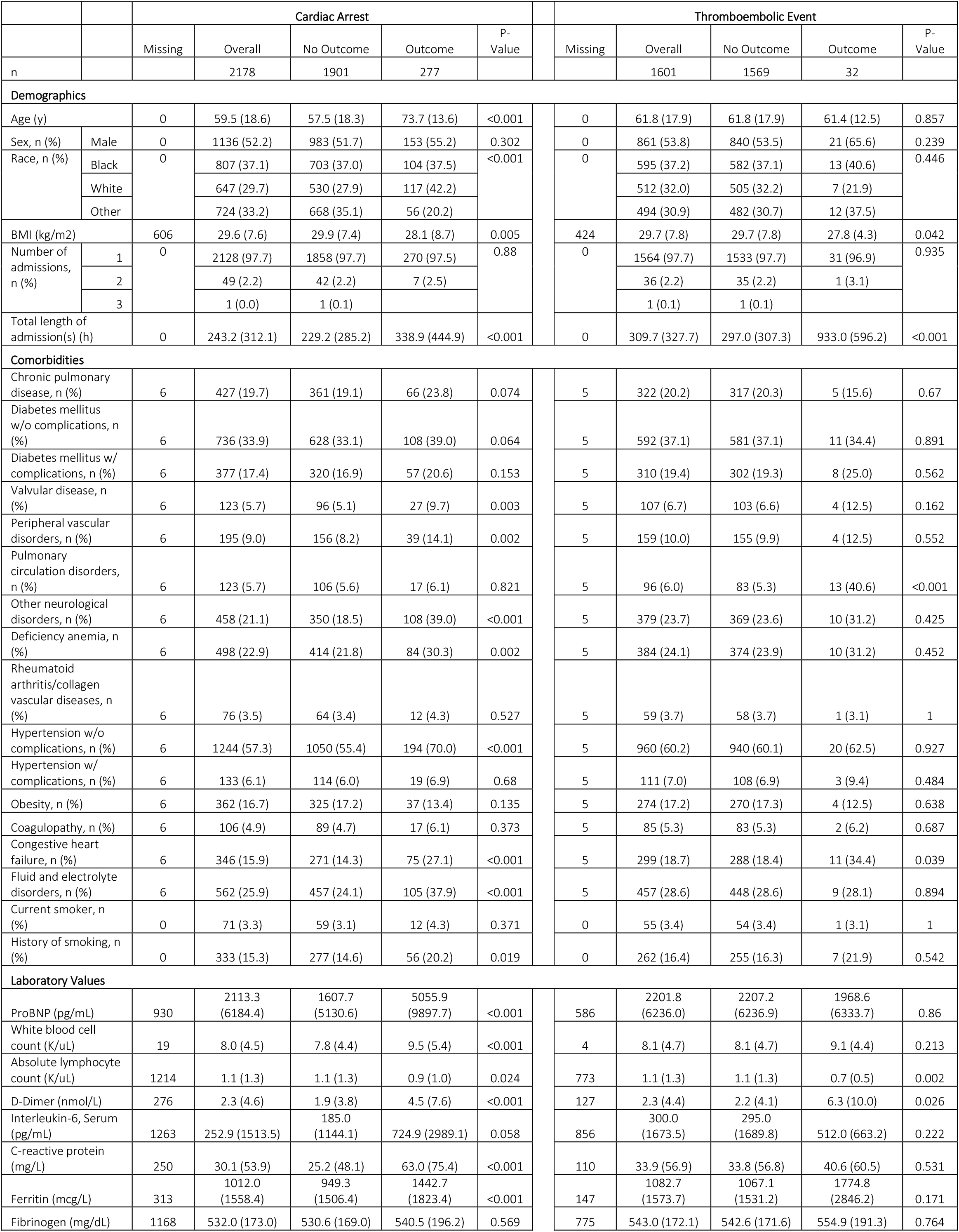
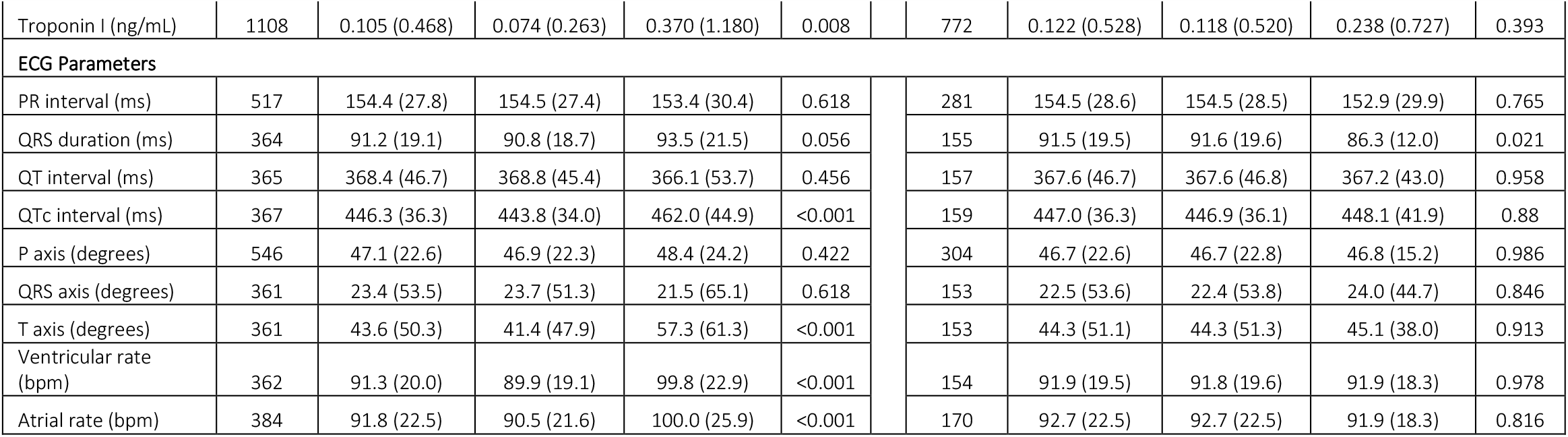
Characteristics of entire dataset for each outcome. ECG parameters and lab values are reported as the first result value during the patient’s admission. Comorbidities are defined according to diagnosis codes in the Elixhauser comorbidity table.^31^ Values are reported mean (standard deviation) unless otherwise indicated. P-values represent comparison between patients that did and did not experience each outcome and were calculated using the two-sample T-test or chi-squared test as appropriate. This table was generated using the python package *tableone* with the Bonferroni correction applied for multiple hypothesis testing.^30^

**Supplementary Table S4:**
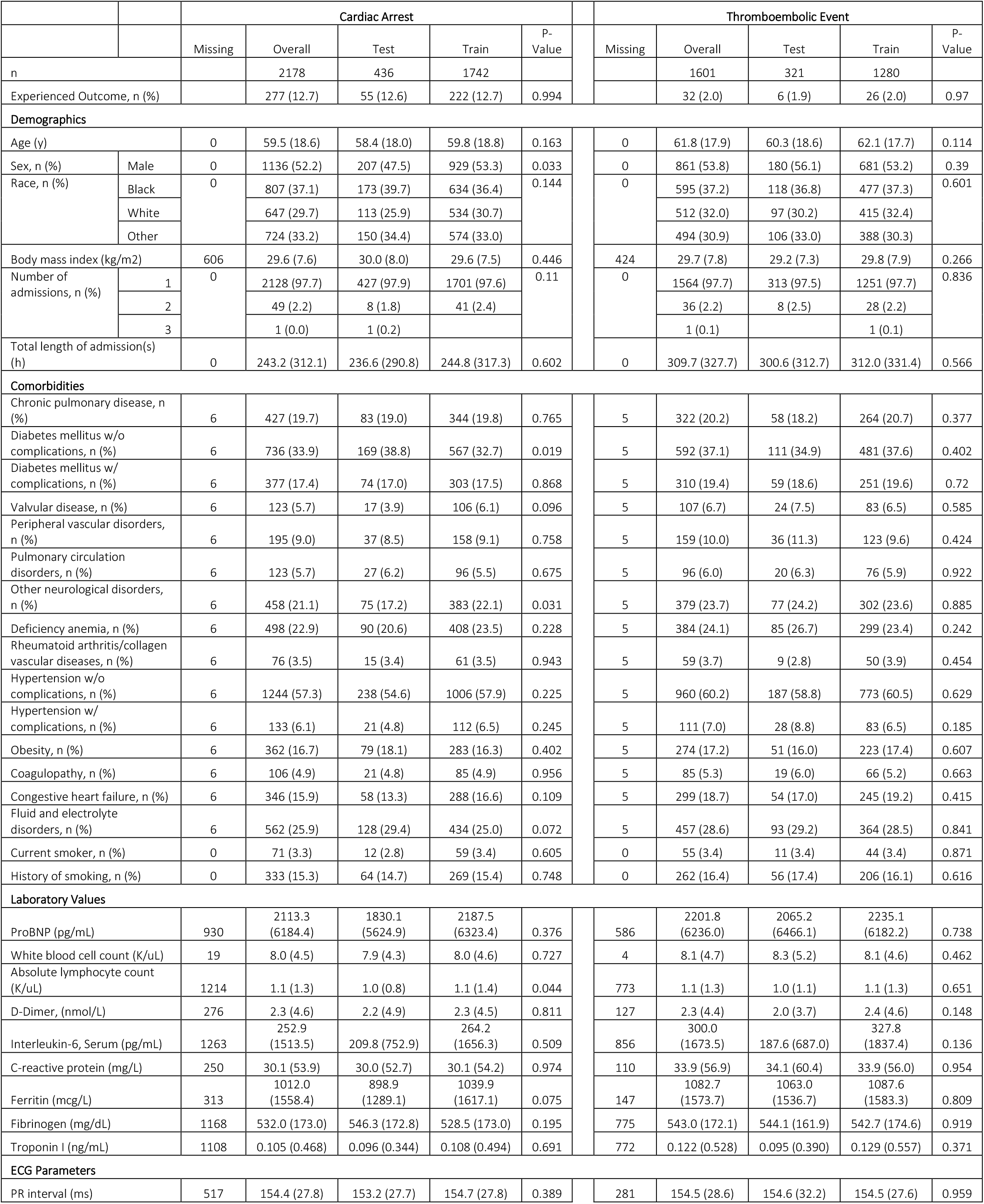
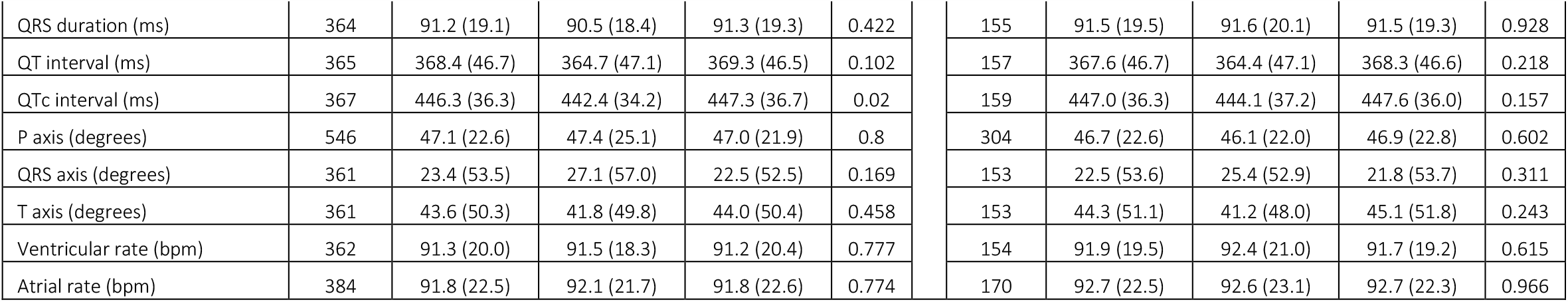
Characteristics of the training and test sets for each outcome. ECG parameters and lab values are reported as the first result value during the patient’s admission. Comorbidities are defined according to diagnosis codes in the Elixhauser comorbidity table.^31^ Values are reported as mean (standard deviation) unless otherwise indicated. P-values represent comparison between patients in the training and test sets for each outcome and were calculated using the two-sample T-test or chi-squared test as appropriate. This table was generated using the python package *tableone* with the Bonferroni correction applied for multiple hypothesis testing.^30^

**Supplementary Figure S1:**
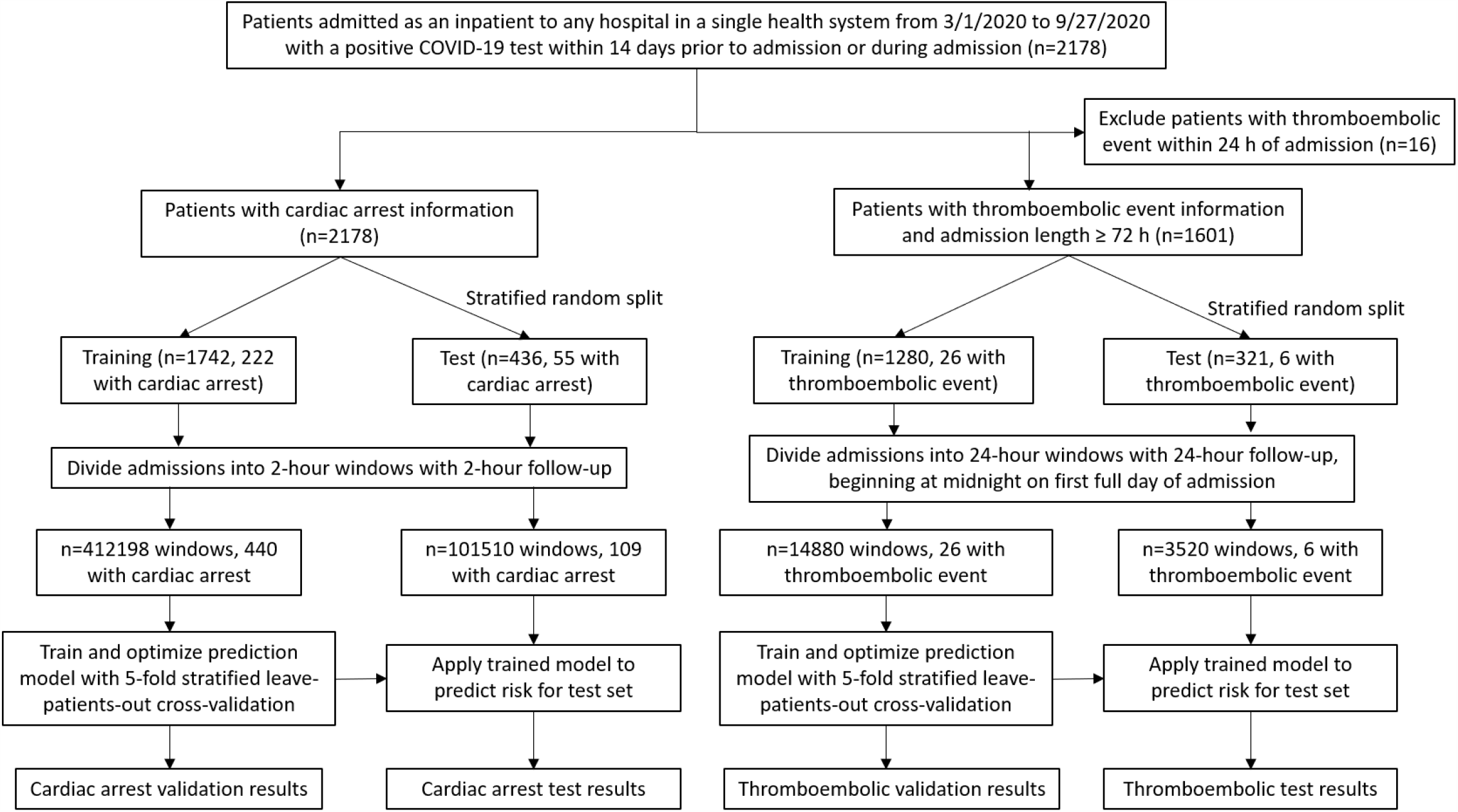
Participant flow diagram for retrospective study of using COVID-HEART to predict cardiac arrest and thromboembolic events continuously over time.

**Supplementary Figure S2:**
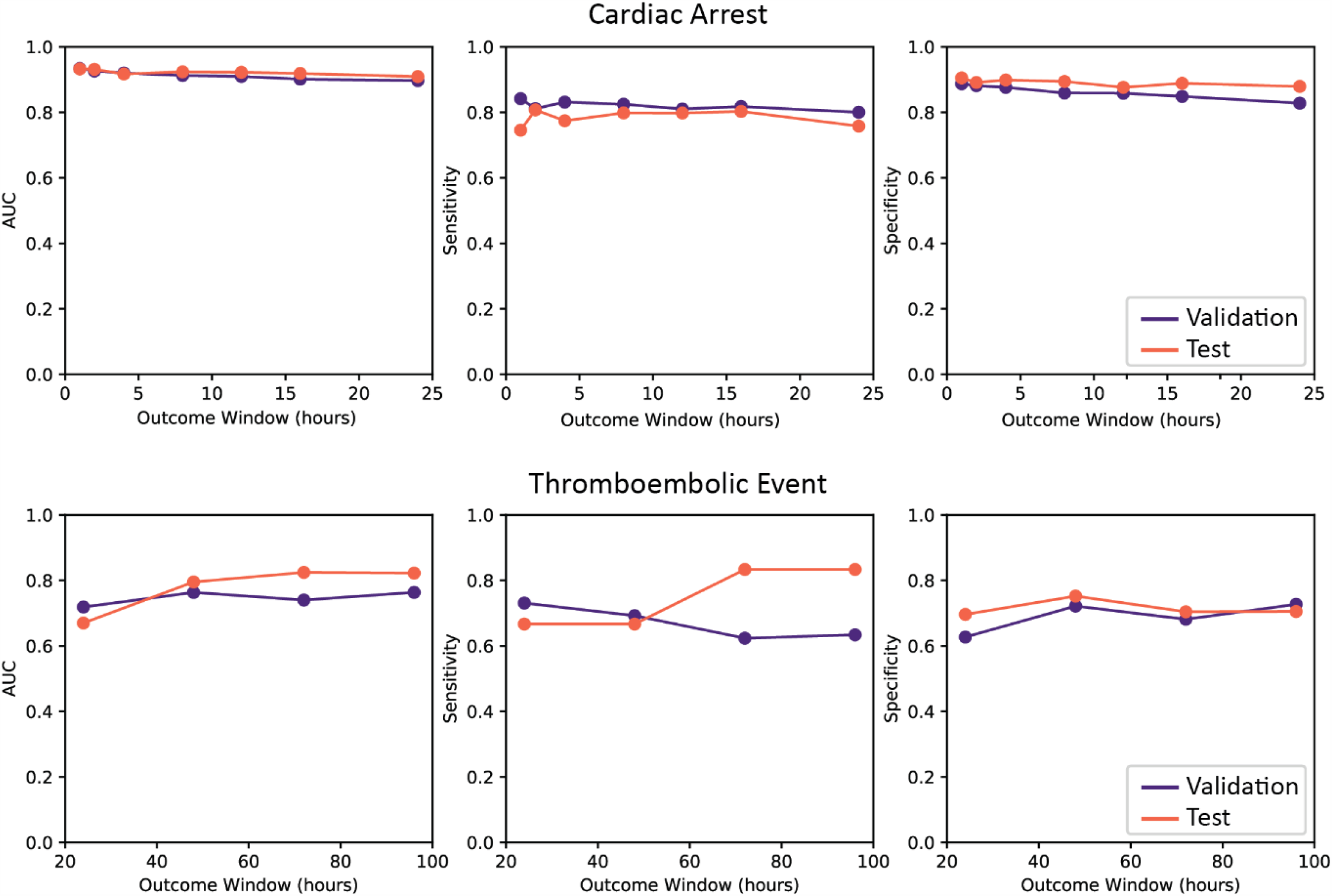
COVID-HEART cross-validation and testing results for outcome windows of different duration in predicting each CV outcome using the optimal classifier. Results for 5-fold stratified patient-based cross-validation (purple) and separate test set (orange) for prediction of cardiac arrest (top) and thromboembolic events (bottom) within a given outcome window using the optimal classifier configuration from Fig.2. Short feature window is 2 hours for prediction of cardiac arrest and 24 hours for prediction of thromboembolic events. Note comparable validation and test results, which indicates strong generalizability. Results shown are for the original development and validation sets (Supplementary Table S2)002E

**Supplementary Figure S3:**
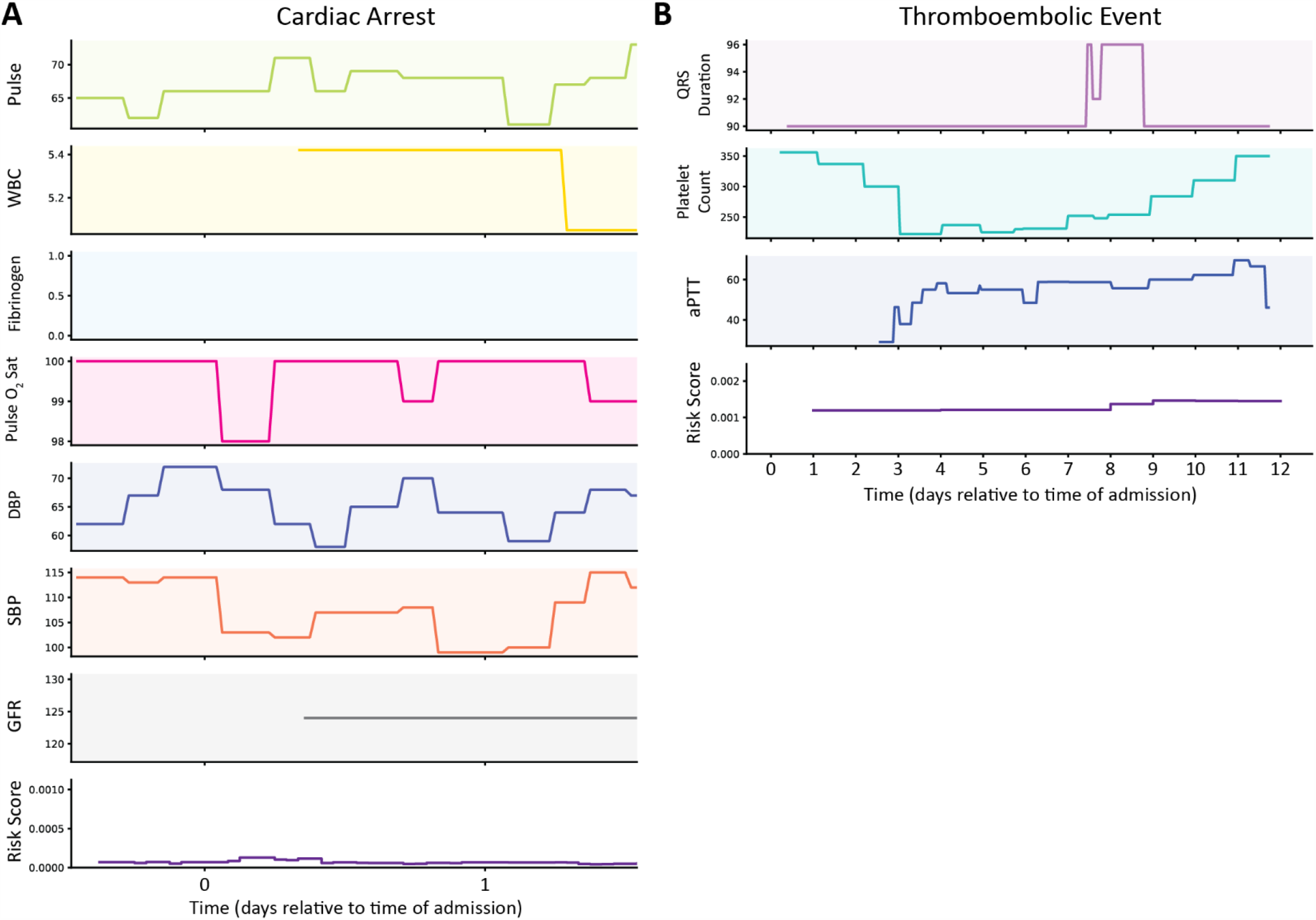
Two examples of “true negative” predictions for two patients, one from the cardiac arrest test set and one from the thromboembolic event test set, using the COVID-HEART predictor. **(A)** Clinical time-series inputs (top 7 rows) from which the features with the largest coefficients were derived for prediction of cardiac arrest and time-series risk score (bottom row) for a patient who did not experience cardiac arrest during their hospitalization, and for whom the classifier’s prediction was correct. The features derived from these inputs are listed in Table 1. A new prediction is generated every hour. The risk score is below 0.1% for the entire duration of the patient’s admission. The date refers to the days of admission relative to midnight on the first full day of admission. Units for each predictor are as follows: Pulse (beats/minutes), WBC (cells/ mm^3^), Fibrinogen (mg/dL), Pulse O_2_ saturation (%), DBP (mmHg), SBP (mmHg), GFR (mL/min) **(B)** Clinical time-series inputs (top 3 rows) from which the 3 selected features were derived for prediction of thromboembolic events, and time-series risk score (bottom row) for a patient who did not experience a thromboembolic event during their hospitalization. The features derived from these inputs are listed in Table 1. A new prediction is generated every 24 hours. The risk score is low for the entire duration of the patient’s admission. The date refers to the days of admission relative to midnight on the first full day of admission. Units for each predictor are as follows: QRS duration (ms), platelet count (cells*10^−3^/uL), aPTT (seconds). *Abbreviations: white blood cell count (WBC), diastolic blood pressure (DBP), systolic blood pressure (SBP), glomerular filtration rate (GFR), activated partial thromboplastin time (aPTT)*

**Supplementary Figure S4:**
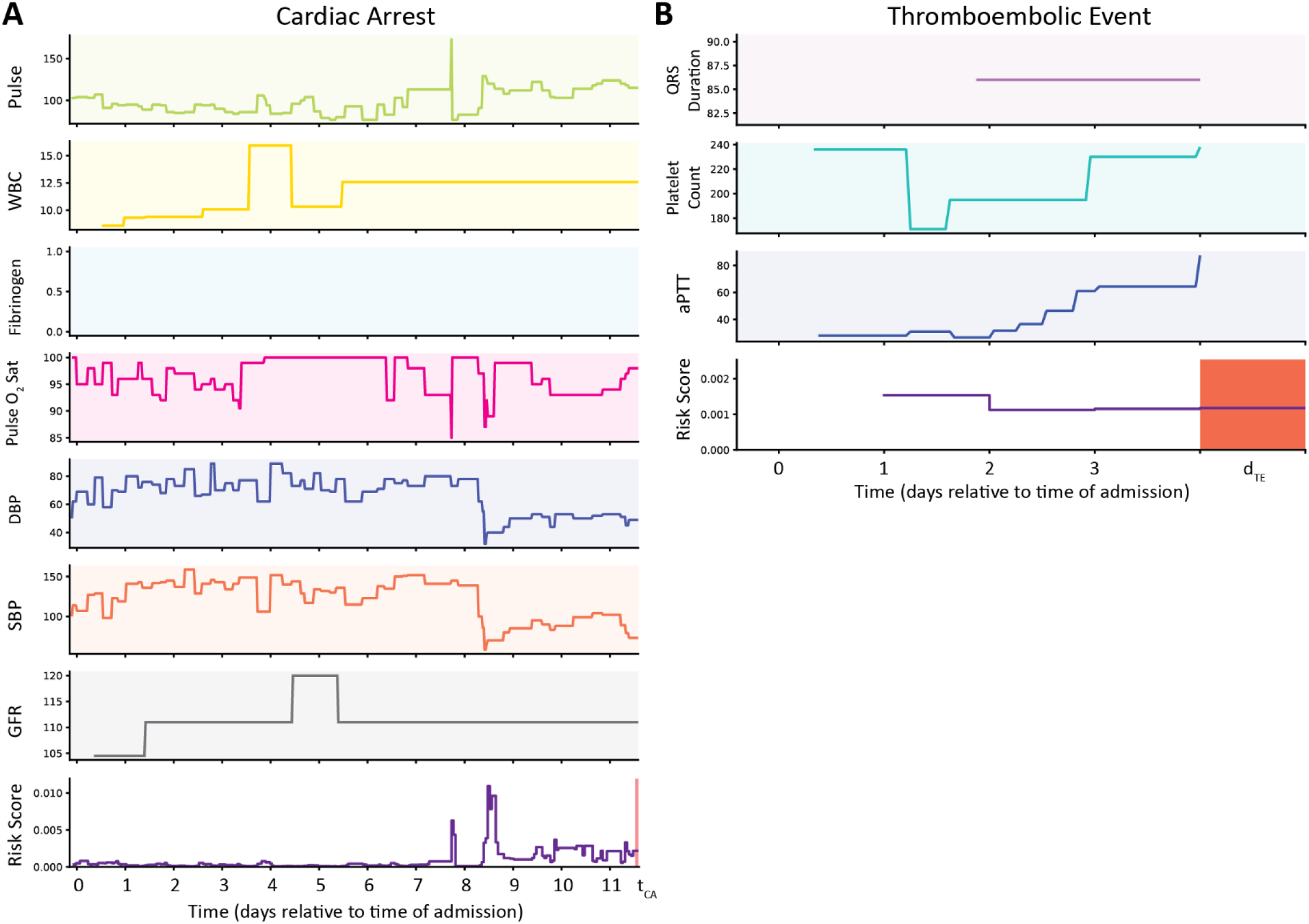
Investigation of incorrect predictions by the COVID-HEART predictor for two patients, one from the cardiac arrest test set and one from the thromboembolic event test set. **(A)** Clinical time-series inputs (top 7 rows) from which the features with the largest coefficients were derived for prediction of cardiac arrest and time-series risk score (bottom row) for a patient who experienced cardiac arrest during their hospitalization, and for whom the classifier’s prediction was correct prior to the cardiac arrest. The features derived from these inputs are listed in Table 1. A new prediction is generated every hour. The risk score is low with slight fluctuations for most of the patient’s hospitalization, then spikes on day 8, then drops but remains above the positivity threshold, which is very low. The patient experienced cardiac arrest on day 11, 3 days after the peak in risk score. The date refers to the days of admission relative to midnight on the first full day of admission. Units for each predictor are as follows: Pulse (beats/minutes), WBC (cells/ mm^3^), Fibrinogen (mg/dL), Pulse O_2_ saturation (%), DBP (mmHg), SBP (mmHg), GFR (mL/min) **(B)** Clinical time-series inputs (top 3 rows) from which the 3 selected features were derived for prediction of thromboembolic events, and time-series risk score (bottom row) for a patient who experienced a thromboembolic event during their hospitalization. The features derived from these inputs are listed in Table 1. A new prediction is generated every 24 hours. The risk score peaks early in the patient’s hospitalization but steadily decreases leading up to the day on which they experienced an imaging-confirmed thromboembolic event; this is discussed in Supplementary Results. The date refers to the days of admission relative to midnight on the first full day of admission. Units for each predictor are as follows: QRS duration (ms), platelet count (cells*10^−3^/uL), aPTT (seconds). *Abbreviations: white blood cell count (WBC), diastolic blood pressure (DBP), systolic blood pressure (SBP), glomerular filtration rate (GFR), activated partial thromboplastin time (aPTT)*

**Supplementary Figure S5:**
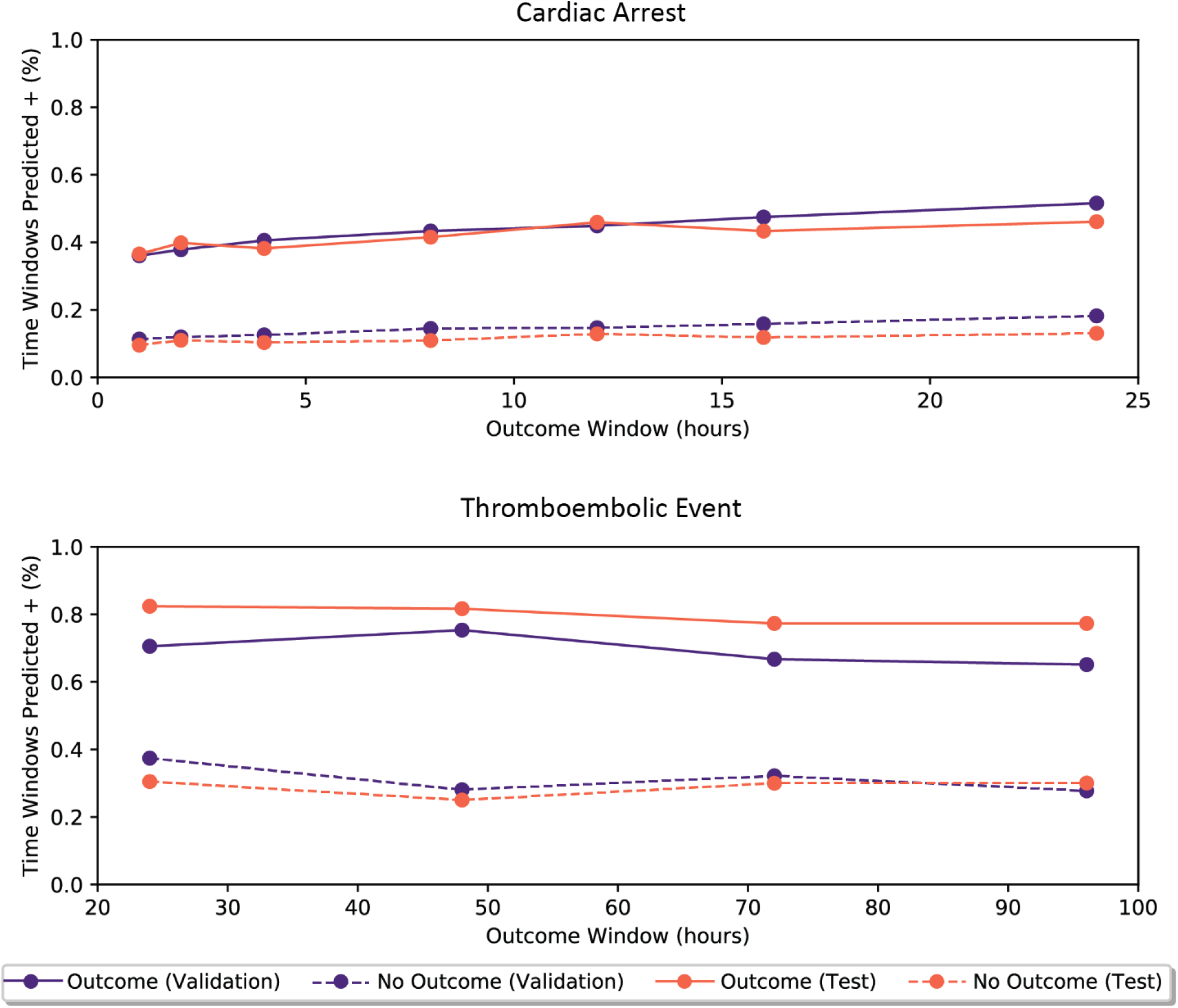
More time windows are predicted positive for patients that eventually experience each outcome than patients who do not. Proportion of time windows predicted positive (risk probability greater than the binary risk threshold determined by the development data) for patients in 5-fold patient-based cross-validation (purple) and the separate test set (orange) that do (solid line) and do not (dashed line) eventually experience cardiac arrest (top) and thromboembolic events (bottom). Results shown are for the original development and validation sets (Supplementary Table S2).

